# SARS-CoV-2 epitopes are recognized by a public and diverse repertoire of human T-cell receptors

**DOI:** 10.1101/2020.05.20.20107813

**Authors:** Alina S. Shomuradova, Murad S. Vagida, Savely A. Sheetikov, Ksenia V. Zornikova, Dmitry Kiryukhin, Aleksei Titov, Iuliia O. Peshkova, Alexandra Khmelevskaya, Dmitry V. Dianov, Maria Malasheva, Anton Shmelev, Yana Serdyuk, Dmitry V. Bagaev, Anastasia Pivnyuk, Dmitrii S. Shcherbinin, Alexandra V. Maleeva, Naina T. Shakirova, Artem Pilunov, Dmitry B. Malko, Ekaterina G. Khamaganova, Bella Biderman, Alexander Ivanov, Mikhail Shugay, Grigory A. Efimov

## Abstract

Understanding the hallmarks of the adaptive immune response to SARS-CoV-2 is critical for fighting the COVID-19 pandemic. We assessed the antibody and T-cell reactivity in COVID-19 convalescent patients and healthy donors sampled both prior to and during the pandemic. The numbers of SARS-CoV-2-specific T cells were increased in healthy donors examined during COVID-19. Combined with the absence of symptoms and humoral response across that group, this finding suggests that some individuals might be protected by T-cell cross-reactivity. In convalescent patients we observed public and diverse T-cell response to SARS-CoV-2 epitopes, revealing T-cell receptor motifs with germline-encoded features. Bulk CD4+ and CD8+ T-cell responses to Spike glycoprotein were mediated by groups of homologous T-cell receptors, some of them shared across multiple donors. Overall, our results demonstrate that T-cell response to SARS-CoV-2, including the identified set of specific T-cell receptors, can serve as a useful biomarker for surveying viral exposure and immunity.

## Introduction

SARS-CoV-2 is currently causing the global pandemic of COVID-19 (Lillie, Samson et al. 2020, Phelan, Katz et al. 2020, Wu, Zhao et al. 2020, Zhou, Yang et al. 2020, Zhu, Zhang et al. 2020). Elucidating the mechanisms of the adaptive immune response to SARS-CoV-2 is crucial for the prediction of vaccine efficacy and assessment of the possibility of reinfection.

It is commonly assumed that the antibody response is required for viral clearance (Huang, Garcia-Carreras et al. 2020). Multiple serological tests for detection of the SARS-CoV-2-specific antibodies are being developed (Amanat, Stadlbauer et al. 2020, Guo, Ren et al. 2020, Krammer and Simon 2020) and massive efforts are undertaken to estimate the number of seropositive individuals in the population. Moreover, monoclonal antibodies (Pinto, Park et al. 2020, Shanmugaraj, Siriwattananon et al. 2020, Wang, Li et al. 2020) and plasma of the convalescent patients were proposed for COVID-19 therapy (Wong, Dai et al. 2003, Chen, Xiong et al. 2020). Administration of monoclonal neutralizing antibodies against glycoprotein S (S-protein) of SARS-CoV-2 protected experimental animals from the high dose of SARS-CoV-2 (Rogers, Zhao et al. 2020). However, around 30% of convalescent patients have no or very low titers of SARS-CoV-2 neutralizing antibodies (Wu, Wang et al. 2020) suggesting the involvement of the other immune mechanisms in the viral elimination.

There is strong evidence of an important role of T-cellular immunity in the clearance of the respiratory viruses, such as SARS-CoV which caused atypical pneumonia outbreak in 2003. Memory T-cell responses to the SARS-CoV epitopes were detectable in 50% of convalescent patients 12 months post-infection (Li, Wu et al. 2008). Moreover, CD8+ cells specific to the immunodominant epitope of S-protein of the mouse-adapted SARS-CoV strain protected aged mice from otherwise lethal infection (Channappanavar, Fett et al. 2014). In another study the adoptive transfer of SARS-CoV-specific CD8+ or CD4+ cells or immunization with peptide-pulsed dendritic vaccine reduced virus titers in the lungs and enhanced survival of mice, thus proving that T cells are sufficient for virus clearance in the absence of antibodies or activation of the innate immune system (Zhao and Perlman 2010). There are studies suggesting a leading role of CD4+ cells in the SARS-CoV clearance. Depletion of the CD4+ T cells at the time of infection delayed the virus clearance whereas depletion of CD8+ T cells had no such effect (Chen, Lau et al. 2010). Furthermore, CD4+ cell response correlated with the positive outcome of SARS-CoV infection (Zhao and Perlman 2010).

In humans, the severe SARS-CoV infection was characterized by the delayed development of the adaptive immune response and prolonged virus clearance (Cameron, Bermejo-Martin et al. 2008). Decreased numbers of T cells strongly correlated with disease severity (Li, Qiu et al. 2004). T cells not only contributed to the resolution of the disease but also formed a long-lasting memory response. The CD8+ and CD4+ cells isolated from the SARS-CoV convalescent patients 4 years after recovery demonstrated IFNγ secretion in response to stimulation by peptide pools derived from S- N- and M-proteins of SARS-CoV (Fan, Huang et al. 2009). SARS-CoV antigen-specific T cells persisted in the convalescent patients for up to 11 years post-infection.

The S-protein was shown to be the most immunogenic among all the SARS-CoV antigens (Li, Wu et al. 2008, Fan, Huang et al. 2009). Moreover, the particular CD4+ and CD8+ T-cell epitopes of SARS-CoV S-protein were identified. Two CD8+ epitopes were presented in HLA-A*02:01 and elicited a specific response in the SARS-CoV convalescent patients but not in healthy donors (Wang, Sin et al. 2004). Additionally, several CD8+ T-cell epitopes were identified and characterized in the M (Yang, Peng et al. 2006, Yang, Peng et al. 2007) and N (Yang, Peng et al. 2006) proteins of SARS-CoV.

At present, growing evidence confirms the importance of T-cell response to the SARS-CoV-2 in disease control. High CD8+ T-cell counts in the lungs correlated with better control of SARS-CoV-2 progression (Liao, Liu et al. 2020). The presence of the T-follicular helper cells and CD8+ T cells with activated phenotype in the blood at the time of virus clearance suggests their active involvement in the immune response in the recovered patient (Thevarajan, Nguyen et al. 2020). On the contrary, the exhausted phenotype of CD8+ T cells in the peripheral blood may serve as an indicator of poor disease prognosis (Zheng, Zhang et al. 2020).

Most probably, the immune memory is capable of protecting against the SARS-CoV-2. The studies on primates demonstrated that the repeated virus challenge failed to provoke reinfection once the virus was eliminated (Bao, Deng et al. 2020).

The T cells of convalescent patients are responsive to stimulation by the peptide pools covering the SARS-CoV-2 proteins. In particular, S-protein of SARS-CoV-2 was a strong inducer of Th1-type response in the CD4+ cells (Weiskopf, Schmitz et al. 2020).

Interestingly the CD4+ cells reactive to the SARS-CoV-2 antigens were found not only in the COVID-19 convalescent patients but also in healthy donors that most likely could be explained by the T-cell cross-reactivity (Braun, Loyal et al. 2020). Further studies are necessary to clarify whether this preexisting T-cell response is protective.

Other studies hinted on the possible role of patient HLA-genotype in the reaction to the virus. According to the bioinformatic predictions, some HLA alleles present more of the SARS-CoV-2 epitopes than the others, possibly affecting the severity of the COVID-19 (Nguyen, David et al. 2020). Some known immunogenic T-cell epitopes of SARS-CoV are conserved in SARS-CoV-2 (Grifoni, Sidney et al. 2020), suggesting that they might also play a role in the immune response to the SARS-CoV-2. However, no experimental data about the targets of T-cell reactivity is currently available.

Thus, we analyzed the adaptive immune response to SARS-COV-2 in the COVID-19 convalescent patients (CP) aiming to describe the underlying structure, clonality, and epitope-specificity of the T-cellular immune response to the SARS-CoV-2 S-protein.

## Results

### SARS-CoV-2-specific T cells but not antibodies are present in healthy donors sampled during COVID-19 pandemic

To study the adaptive immune response to SARS-CoV-2 we recruited 31 COVID-19 convalescent patients (CP). According to the classification by the US National Institutes of Health (https://covid19treatmentguidelines.nih.gov/overview/management-of-covid-19/) the patients were categorized as having asymptomatic (n = 2), mild (n = 19) or moderate/severe (n = 10) clinical types of disease. None of the patients required either treatment in the intensive care unit, oxygen supplementation, or invasive ventilation support. The cohort was gender-balanced (15 males, 16 females). Patient age was from 17 to 59 years with a median of 35 years. Peripheral blood was collected at days 17 - 49 (median 33) after the onset of the disease or positive PCR-test (Figure 1a). The control group of healthy donors was formed by recruiting 14 healthy volunteers during the COVID-19 pandemic - HD (CoV) with no symptoms and a negative PCR test. Additionally, we used 10 samples of peripheral blood mononuclear cells (PBMC) from biobanked healthy hematopoietic stem cell donors - HD (BB), which were cryopreserved no later than September 2019, and 10 serum samples of healthy blood donors cryopreserved no later than 2017 - HD (S). The presence of SARS-CoV-2 specific antibodies was tested in the serum samples. Cell samples were used to study T-cellular response to the peptide pools of M-, N-, S-proteins of SARS CoV-2, and to the recombinant surface glycoprotein S (S-protein) as well as to determine the repertoire of T-cell receptors (TCRs) of S-protein specific cells (Figure 1b).

**Figure 1.**
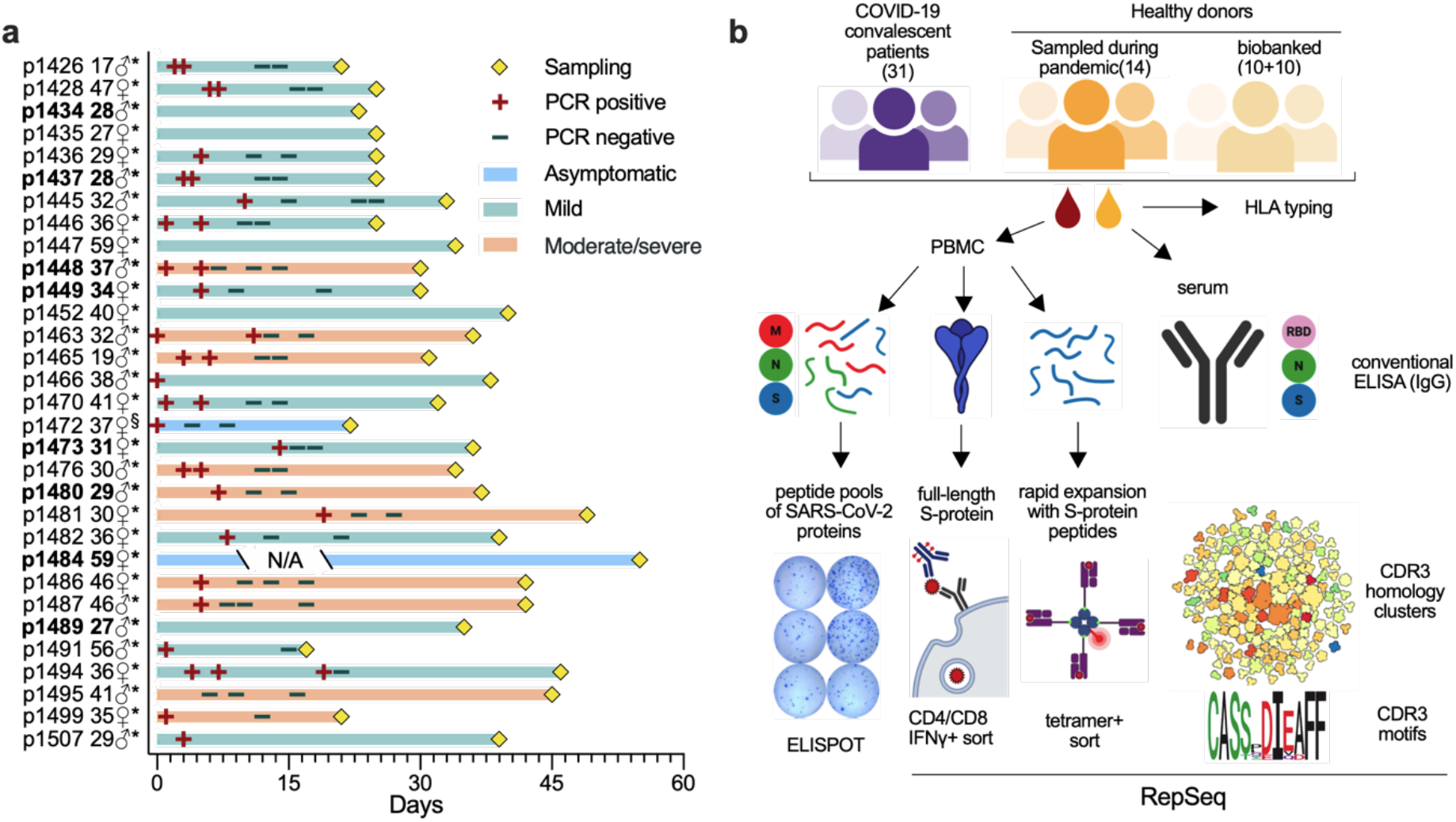
Experimental pipeline and COVID-19 patient data. **(a)** Clinical data of COVID-19 CP. Age and gender are indicated on the left of the Y-axis. The time point of sampling, results of PCR tests and severity of disease are provided on the swimmer’s plot. * - days are calculated since the onset of symptoms, § - days are calculated since the positive PCR test. N/A - information is not available. Donors for which the TCR repertoire was analyzed are shown in bold. **(b)** The cohorts of COVID-19 convalescent patients - CP (n=31), healthy donors sampled during COVID-19 pandemic - HD(CoV) (n=14) and obtained from the biobank - HD(BB) (n=10) and serum bank - HD(S) (n=10) were included in study. Peripheral blood was collected and used for in vitro assays with several SARS-CoV-2 antigens. Antigen-specific IFNγ production by T cells in CP to S-protein and peptide pools of SARS-CoV-2 (M, N, and S) was measured by ELISPOT. The S-protein-directed T-cell response was assessed by IFNγ secretion assay after stimulation of peripheral blood mononuclear cells (PBMC) with recombinant S-protein, FACS-sorted IFNγ-secreting CD4+ and CD8+ T-cells were used for sequencing their T-cell receptor (TCR) beta repertoire. Simultaneously, rapid in vitro expansions with predicted HLA-A*02:01-presented S-protein derived peptides were set up, tetramer-positive cells were FACS-sorted and TCR alpha and beta repertoire sequencing was performed. Bioinformatic analysis was used to reveal shared CDR3 motifs in antigen and epitope-specific populations. Antibody-mediated immune response in CP and HD was measured by conventional ELISA with recombinant N-, S-protein, and RBD domain of S-protein. Illustration was created with BioRender.

Analysis of the humoral immune response to SARS-CoV-2 demonstrated that the majority of COVID-19 CPs had IgG antibodies specific to all of the tested viral antigens. IgGs from HD (CoV) and HD (BB) groups showed no reactivity to S-protein of SARS-CoV-2 or its receptor-binding domain (RBD) (Figure 2a and 2b). The presence of antibodies specific to the nucleoprotein (N-protein) of SARS-CoV-2 in HD could be explained by either their cross-reactivity or by recognition of some bacterial products co-purified with the antigen (Figure 2a and S1a).

**Figure 2.**
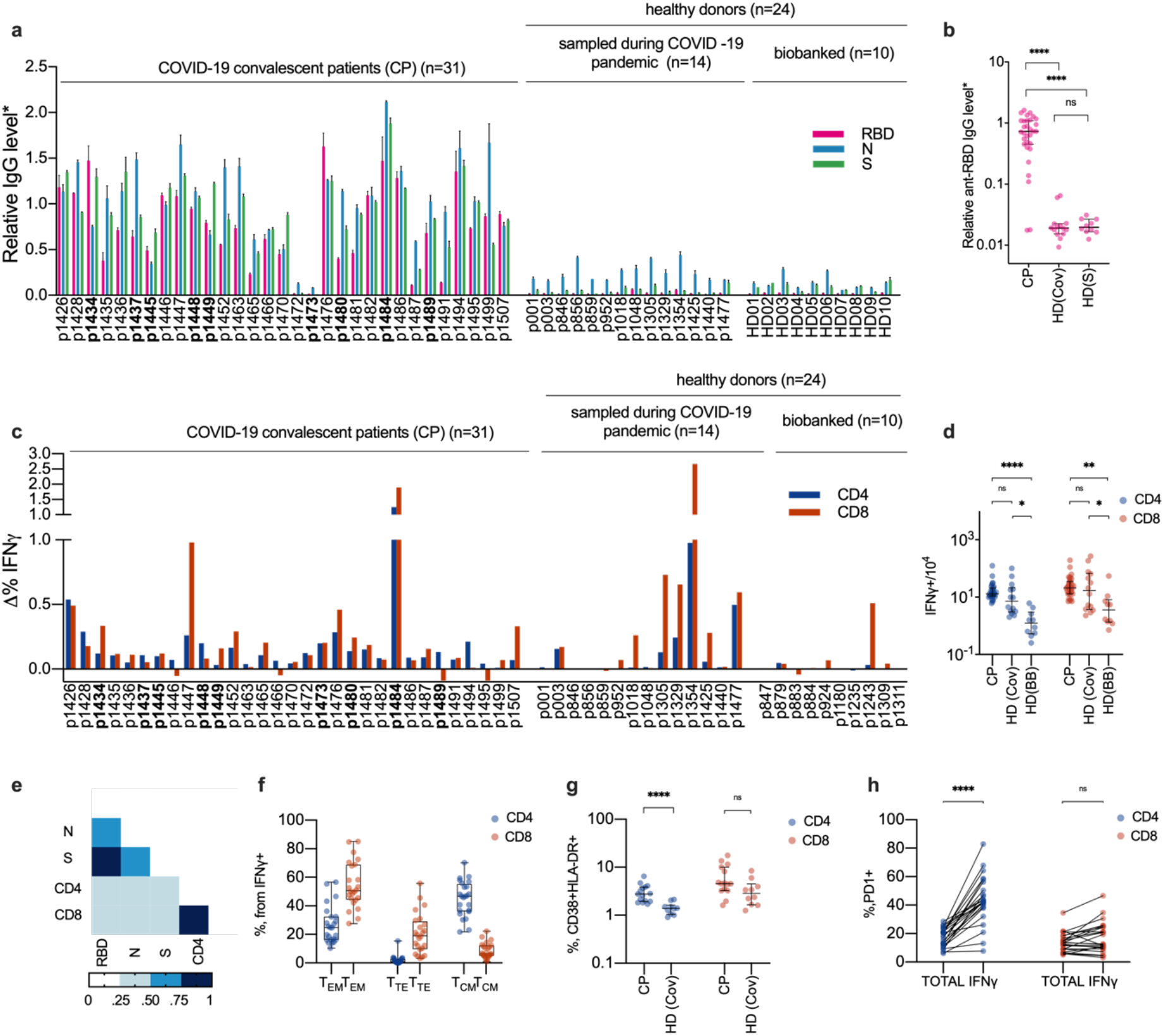
Healthy donors sampled during the COVID-19 pandemic have increased numbers of SARS-CoV-2-specific T cells but not antibodies. **(a-b)** Relative levels of anti-RBD, anti-N and anti-S IgG were measured by conventional ELISA in COVID-19 CP (n=31), HD (CoV) (n=14) and HD (S) (n=10). Plotted data are mean of two independent measurements ± SD (**a**) and medians with bars representing interquartile range **(b)**. **(c-d)**, T-cell response to S-protein was measured by frequencies of IFNγ-producing CD4+ and CD8+ cells in COVID-19 CP (n=31), HD (CoV) (n=14) and HD (BB) (n=10). Data are delta between frequencies of activated T-cells in stimulated and unstimulated samples **(c)** and medians of the frequencies with bars represent the interquartile range **(d)**. **(e)** Correlation between relative levels of antibodies and T-cell response in CP group (n=31). Heat-map of intervals of spearman’s coefficients of correlation is shown. **(f)** Phenotype of IFNγ-secreting CD4+ and cD8+ cells in CP (n=24). TEM - T effector memory (CD45RO+, CD197-), TTE - T terminal effector (CD45RO-, CD197-) and TCM - T central memory (CD45RO+, CD197+). Box represents interquartile range with the median line, whiskers represent min and max. (n=24). **(g)** Percentage of activated (CD38+ HLA-DR+) CD4+ and CD8+ cells in CP (n=15) and HD (CoV) (n=10) groups. Data are median with interquartile range. **(h)** Comparison of frequencies between PD1 + cells in CD4+ and CD8+ not-naive T cells (TOTAL) and not-naive activated subpopulation (IFNγ) in CP group (n=24). For group comparison, we used the Kruskal-Wallis test and Dunn’s multiple comparison test **(b-d)** and Mann-Whitney test **(g-h**). p-value *p<0.05, **p<0.01, ***p<0.001, ****p<0.0001. CP - convalescent patients, HD (CoV) - healthy donors sampled during COVID-19 pandemic, HD (BB), and HD (S) - biobanked healthy donor cell or serum samples, respectively.

Despite the variability of the antibody response, levels of IgG specific to all three antigens in most cases distinguished CP from HD (Figure 2a, 2b and S1a, S1b), while using RBD resulted in the lowest background. Levels of IgG antibodies specific to different antigens positively correlated in CP (Figure 2e and S1c-e) with the strongest correlation (r=0.82, p<0.0001) between RBD and S-protein. Only two patients (p1472 and p1473) did not demonstrate IgG response to any of the tested viral antigens.

T-cell response to the SARS-CoV-2 S-protein was highly variable across the donors with some CP lacking detectable virus-reactive T cells (Figure 2c and 2d). We have not observed any clear association between the magnitude of T-cell response and the HLA genotype of the donor (Table S1), time after the disease onset, its severity or patient age (Figure S2j-o). We observed only mild correlation between the magnitude of T-cell and humoral response in our cohort (for anti-RBD IgG and CD8+ T-cell response r=0.386 p=0.0321) whereas the magnitude of CD8+ and CD4+ responses were interdependent (Figure 2e).

All tested HD(CoV) lacked antibodies against any of SARS-CoV-2 antigens, which probably excludes the possibility of past asymptomatic infection. Surprisingly some of them had frequencies of S-protein specific T-cells comparable with those in the CP group (Figure 2c and 2d). Apart from the significant difference in T-cell response between CP and HD(BB) (for CD4+ p<0.0001; for CD8+ p=0.0014), we also observed the increase of both CD4+ and CD8+ S-protein specific T-cells in HD(CoV) as compared with HD(BB) group (for CD4+ p=0.0106; for CD8+ p=0.0456) (Figure 2d and S3). This might indicate that some HD(CoV) were exposed to the virus but did not develop the disease.

S-protein specific T cells in CP exhibited conventional phenotype distribution typical to CD4+ and CD8+ cells. S-protein reactive CD4+ lymphocytes were represented predominantly by central memory phenotype (CD45RO+, CD197+), followed by the effector memory phenotype (CD45RO+, CD197-). Antigen-specific CD8+ cells had mostly effector memory phenotype, followed by the terminal effector cells (CD45RO-, CD197-) (Figure 2f and S3). Besides, we observed a significant increase of activated (CD38+, HLA-DR+) CD4+ cells in the CP group compared to HD(Cov) (Figure 2g and S3). The level of PD-1 expression by CD4+ but not CD8+ cells was significantly higher in the IFNy-secreting population (Figure 2h). Flow cytometry gating strategy for all population is shown on the Figure S3.

We also measured T-cell immune response to the recombinant SARS-CoV-2 S-protein using ELISPOT and to the peptide pools covering SARS-CoV-2 S, M, and N-proteins. Some patients responded to recombinant S-protein while they demonstrated no response to S-protein derived peptide pools (Figure S4). This might be explained by the incomplete coverage of the protein sequence (refer to the discussion for details). Activation of T cells upon full-length S-protein stimulation was equally effective in both CD4+ and CD8+ lymphocytes (Figure S4a and S4b) The response to M-protein was significantly stronger (p=0.0352) (Figure S4c and S4d).

All CP exhibited either CD8+ or CD4+ T-cell reactivity to at least one of the proteins of SARS CoV-2 (Figure 2c, 2d and S4).

### Immune response to two HLA-A*02:01-restricted SARS-CoV-2 S-protein epitopes allowed to discriminate convalescent patients from healthy donors

The most common MHC I allele in the CP cohort was HLA-A*02:01 (Table S1) present in 14 of the 31 patients. We selected 13 potential S-protein epitopes predicted to be presented by HLA-A*02:01, some of them shared 100% sequence homology with SARS-CoV and were previously shown to be immunogenic (Table 1). Total magnitude of S-protein directed CD8+ response was in some patients less than 0.1% from the total CD8+ population, so we decided to perform rapid in vitro antigen-specific expansion according to the protocol which was previously proposed to expand antigen-specific cells from the memory subpopulation (Danilova, Anagnostou et al. 2018). Epitope-specific cells were detected by flow cytometry using MHC-tetramers (Figure 3a and 3b). Strikingly, of 14 CPs with HLA-A*02:01 allele reacted to a single epitope of S-protein_269-277_ YLQPRTFLL (YLQ), while only 1 HD(Cov) had the cells recognizing this epitope. The response to the second epitope S-protein_1000-1008_ RLQSLQTYV (RLQ) was also, though to a somewhat lesser extent, pointing to the past COVID-19 (Figure 3a). A proportion of the positive wells in expansion for both epitopes could be used to confirm previous COVID-19 (Figure 3c). Almost complete absence of the response to these epitopes in HD confirmed that the detected antigen-specific cells were derived from the memory subpopulation. The remaining 11 of the tested epitopes yielded only sporadic T-cell response (Figure 3a).

**Table 1.** Set of HLA-A*02:01-restricted peptides of S-protein used in this study.

| N | Start position | End position | Length | AA sequence | HLA restriction | Cleavage probability | Binding Score | Binding Rank | SARS-CoV-1 identity | Link |
| --- | --- | --- | --- | --- | --- | --- | --- | --- | --- | --- |
| 1 | 269 | 277 | 9 | YLQPRTFLL | A*02:01 | 0.977383 | 0.973032 | 0.015 | 6/9 | (Baruah and Bose 2020) |
| 2 | 417 | 425 | 9 | KIADYNYKL | A*02:01 | 0.966616 | 0.908998 | 0.0583 | 8/9 | - |
| 3 | 424 | 433 | 10 | KLPDDFTGCV | A*02:01 | 0.896237 | 0.591582 | 0.3676 | 8/10 | (Grifoni, Sidney et al. 2020) |
| 4 | 691 | 699 | 9 | SIIAYTMSL | A*02:01 | 0.951629 | 799,513 | 0,11 | 8/9 | (Lv, Ruan et al. 2009) |
| 5 | 821 | 829 | 9 | LLFNKVTLA | A*02:01 | 0.859019 | 0.785743 | 0.1599 | 9/9 | (Ishizuka, Grebe et al. 2009) |
| 6 | 958 | 966 | 9 | ALNTLVKQL | A*02:01 | 0.943025 | 0.450592 | 0.6159 | 9/9 | (Grifoni, Sidney et al. 2020) |
| 7 | 976 | 984 | 9 | VLNDILSRL | A*02:01 | 0.97061 | 0.950712 | 0.0356 | 9/9 | (Lv, Ruan et al. 2009) |
| 8 | 983 | 991 | 9 | RLDKVEAEV | A*02:01 | 0.969105 | 0.860941 | 0.0962 | 9/9 | - |
| 9 | 996 | 1004 | 9 | LITGRLQSL | A*02:01 | 0.891839 | 0.957880 | 1.98 | 9/9 | (Wang, Sin et al. 2004) |
| 10 | 1000 | 1008 | 9 | RLQSLQTYV | A*02:01 | 0.748401 | 0.743108 | 0.199 | 9/9 | (Wang, Chen et al. 2004) |
| 11 | 1185 | 1193 | 9 | RLNEVAKNL | A*02:01 | 0.9655 | 0.618928 | 0.3295 | 9/9 | (Wang, Chen et al. 2004) |
| 12 | 1192 | 1200 | 9 | NLNESLIDL | A*02:01 | 0.945978 | 0.697243 | 0.2411 | 9/9 | (Lv, Ruan et al. 2009) |
| 13 | 1220 | 1228 | 9 | FIAGLIAIV | A*02:01 | 0.179154 | 0.82072 | 0.1288 | 9/9 | (Wang, Sin et al. 2004) |

**Figure 3.**
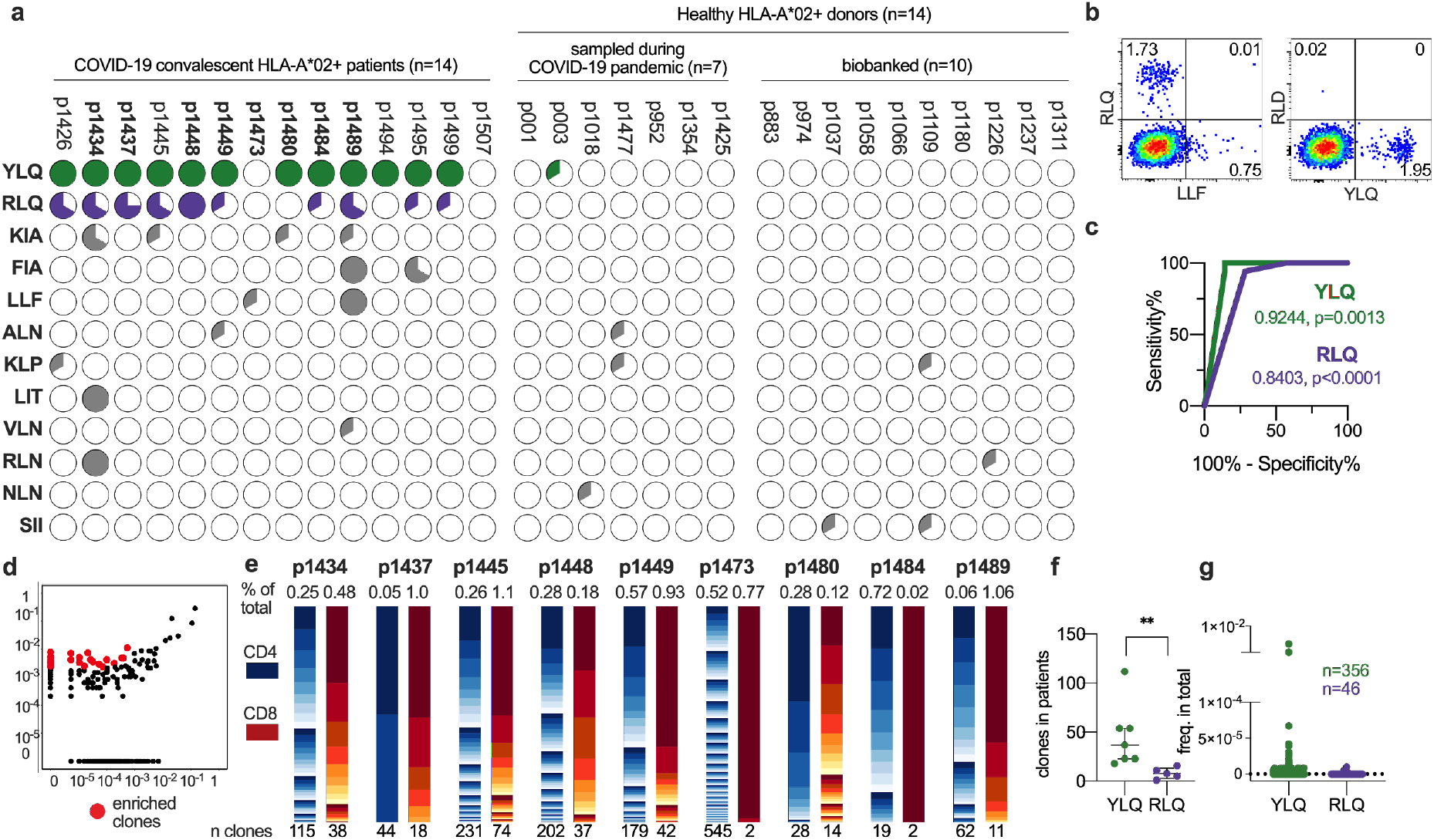
Epitope specificity of CD8+ T-cell response to SARS-CoV-2 S-protein is significantly different between HLA-A*02+ COVID-19 convalescent patients and healthy donors. **(a-b)** pbmc of HLA-A*02:01 positive CP (n=14), HD(Cov) (n=7) and HD(BB) (n=10) were stimulated with a mix of 13 predicted peptides and expanded for 8-12 days followed by MHC-tetramer staining. **(a)** Pie charts represent fractions of wells containing tetramer-positive cells after expansion. **(b)** Representative MHC-tetramer staining at day 9 (patient p1445, well 3). **(c)** ROC curve of the CP/HD classifier based on the presence of YLQ- or RLQ-epitope-specific cells after expansion. **(d-e)** The IFNγ-secreting CD4+ and CD8+ cells were FACS-sorted after stimulation with the S-protein and their TCR beta repertoire was sequenced. **(d)** Representative enrichment plot is shown (patient p1448, IFNγ-secreting CD8+ vs total PBMC). Red dots represent strongly (>10X) and significantly (p<10^-8^, exact Fisher test) enriched clones, assumed to be antigen-specific. **(e)** Clonal structure of the CD4+ (blue) and CD8+ (orange) antigen-specific T-cell population. Numbers below the bars indicate the total number of antigen-specific clones, numbers above - their combined share in the total T-cell repertoire. **(f)** MHC-tetramer-positive clones after rapid in vitro expansion were FACS sorted and their TCR repertoires were sequenced. Numbers of YLQ- and RLQ-specific clones in each patient are plotted (p=0.0013 by Mann-Whitney test). **(g)**YLQ-and RLQ-specific T cell clones occupy only a negligible fraction of the total T cells repertoire. Frequencies of each antigen-specific T cell clone in the PBMC are plotted.

To describe the structure and clonality of SARS-CoV-2 directed T-cell immune response we performed the analysis of T-cell receptor (TCR) repertoires of FACS-sorted IFNγ-secreting CD8+/CD4+ cells and MHC-tetramer-positive populations as well as the total fraction of PBMC by high throughput sequencing using the Illumina platform.

Antigen and epitope-specific TCRs were defined as being strongly and significantly enriched in the respective fraction (Figure 3d).

Clonality of IFNy-secreting cells was higher in CD4+ T cells (Figure 3e) with the number of IFNy-secreting CD4+ clones ranging between 19 and 545 (median=115). The CD8+ IFNy-secreting cell population had a median of 18 (2 to 74) clones (Figure 3e). Sequences of YLQ- and RLQ-specific clonotypes were deposited to the VDJdb database (vdjdb.cdr3.net). Clonotypes significantly enriched in the IFNy-secreting CD4+ and CD8+ populations are listed in Table S2.

We observed only a negligible intersection between MHC-tetramer-positive and IFNγ-secreting populations, indicating only a minimal presence of RLQ- and YLQ-specific clones in the peripheral blood. The YLQ-specific response was significantly more diverse than the RLQ-specific clones with a median of 37 and 8 clones per individual, respectively (Figure 3f). T-cell clones specific for both antigens were either undetectable or observed with a very low frequency in the total TCR repertoire of the peripheral blood (Figure 3g).

### TCRs specific to two SARS-CoV-2 epitopes display prominent sequence motifs shared across individuals

Statistical analysis of V(D)J rearrangement properties of RLQ- and YLQ-specific TCRs revealed biases in the CDR3 length distribution (Figure 4a) and Variable gene usage (Figure 4b) for both TCR alpha (TRA) and beta (TRB) chains. CDR3 regions of these TCRs, with the exception of RLQ-specific TRB, appear to be substantially shorter than those observed in a control PBMC repertoire, a feature previously shown to be associated with “public” TCRs that have higher V(D)J rearrangement probability and incidence rate across individuals (Pogorelyy, Fedorova et al. 2018). TRB CDR3 length difference can also explain why YLQ-specific T-cells are more frequent than RLQ-specific ones.

**Figure 4.**
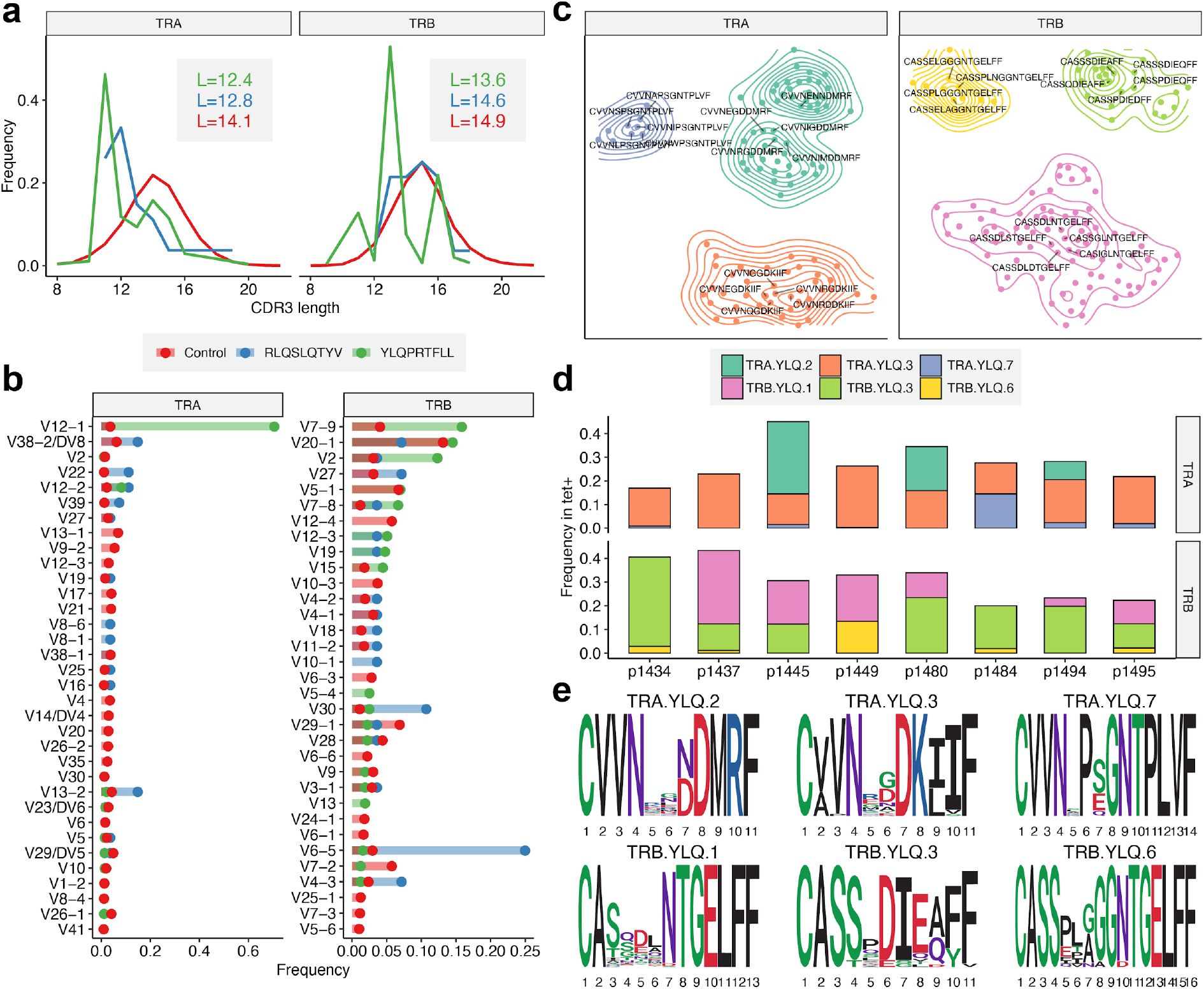
YLQ-specific clones display prominent CDR3 motifs that are shared across individuals. **(a)** Length distribution of CDR3 amino acid sequences of tetramer-positive T cell repertoires and control (background) dataset. Line color indicates the dataset: blue-TCRs specific to epitopes RLQ, green-TCRs specific to epitopes YLQ, red-control PBMC TCR repertoire, L-mean CDR3 lengths The insert shows mean CDR3 lengths. **(b)** A histogram of Variable gene usage across datasets. Only Variable genes with frequency not less than 1 % in any of the datasets are shown. **(c)** CDR3 sequence similarity maps of TCR motifs discovered in TCR alpha (TRA) and beta (TRB) chain tetramer-positive T cell repertoires. Sets of highly homologous CDR3 sequences with at least ten members are shown, contour plots show connected components of corresponding sequence similarity graph, labels highlight five - representative CDR3 sequences for each cluster. **(d)** Distribution of TCRs corresponding to each CDR3 motif across donor samples. The color of each bar corresponds to specified CDR3 motif. Height of each colored bar corresponds to the total fraction of T-cells having a given CDR3 in each donor. **(e)** Position-weight matrices (PWMs) for CDR3 sequences of TRA and TRB chain motifs found in tetramer-positive T-cell repertoires.

We observed some notable differences in the frequency of certain Variable genes: for example, TRAV12-1 and TRBV7-9 were used by 71% and 16% YLQ-specific TCRs, TRAV13-2 and TRBV6-5 were used by 15% and 25% RLQ-specific TCRs, compared to just 3-4% gene usage in control TCRs. Strong biases observed for gene usage might suggest the importance of germline-encoded features in TCR recognition of RLQ and YLQ epitopes and is reminiscent of previously reported TRAV12 bias for the Yellow fever virus LLW epitope (Bovay, Zoete et al. 2018, Minervina, Pogorelyy et al. 2020).

We performed TCR sequence similarity analysis to extract the set of motifs that governs the recognition of RLQ and YLQ epitopes (see Materials and Methods). Our analysis revealed a set of three distinct CDR3 alpha and three distinct CDR3 beta motifs, each containing more than 10 highly similar sequences coming from YLQ-specific T-cells (Figure 4c). Interestingly, all of the motifs were encountered in most of the donors surveyed (Figure 4d), suggesting the public nature of the response and little difference between motifs in terms of publicity. Moreover, some YLQ-specific TCRs were shared between multiple individuals and others exhibited a high degree of global similarity (Figure S5). No significant correlation in frequency was observed between pairs of TRA and TRB motifs, suggesting that the pairing between motifs of different chains may be entirely random, in line with recent observations (Shcherbinin, Belousov et al. 2020). Position-weight matrices of the motifs demonstrate a set of highly dissimilar consensuses (Figure 4e) suggesting that while RLQ- and YLQ-specific TCR repertoires are highly public, they are also diverse.

### TCR repertoire structure analysis of CD4+ and CD8+ T-cell responses to SARS-CoV-2 S-protein reveals public TCR motifs

We analyzed the repertoire sequencing data for bulk T-cell responses to the full-length S**-** protein of SARS-CoV-2 using TCRNET algorithm (see Materials and Methods) that was successfully employed for similar tasks previously (Ritvo, Saadawi et al. 2018). This algorithm detects groups of homologous TCR sequences that are unlikely to arise due to convergent V(D)J recombination and thus are a hallmark of an antigen-specific response. We scored each TCR sequence in a pooled dataset of IFNγ-producing CD4+ and CD8+ T cells by quantifying the number of similar TCR sequences observed in the same dataset and in the control PBMC dataset (Figure 5a). This analysis revealed 732 and 517 unique TRB clonotypes that are part of homologous TCR clusters for CD4+ and CD8+ subsets, respectively (Table S3). Of them, 433 of CD4+ and 195 of CD8+ TRB clonotypes overlapped with those determined above based on the increased clonotype abundance in stimulated fraction compared to control. Only two CDR3 amino acid sequences of tetramer-positive T-cells werematched to these clusters when allowing a single amino acid substitution, both coming from YLQ-specific T-cells.

**Figure 5.**
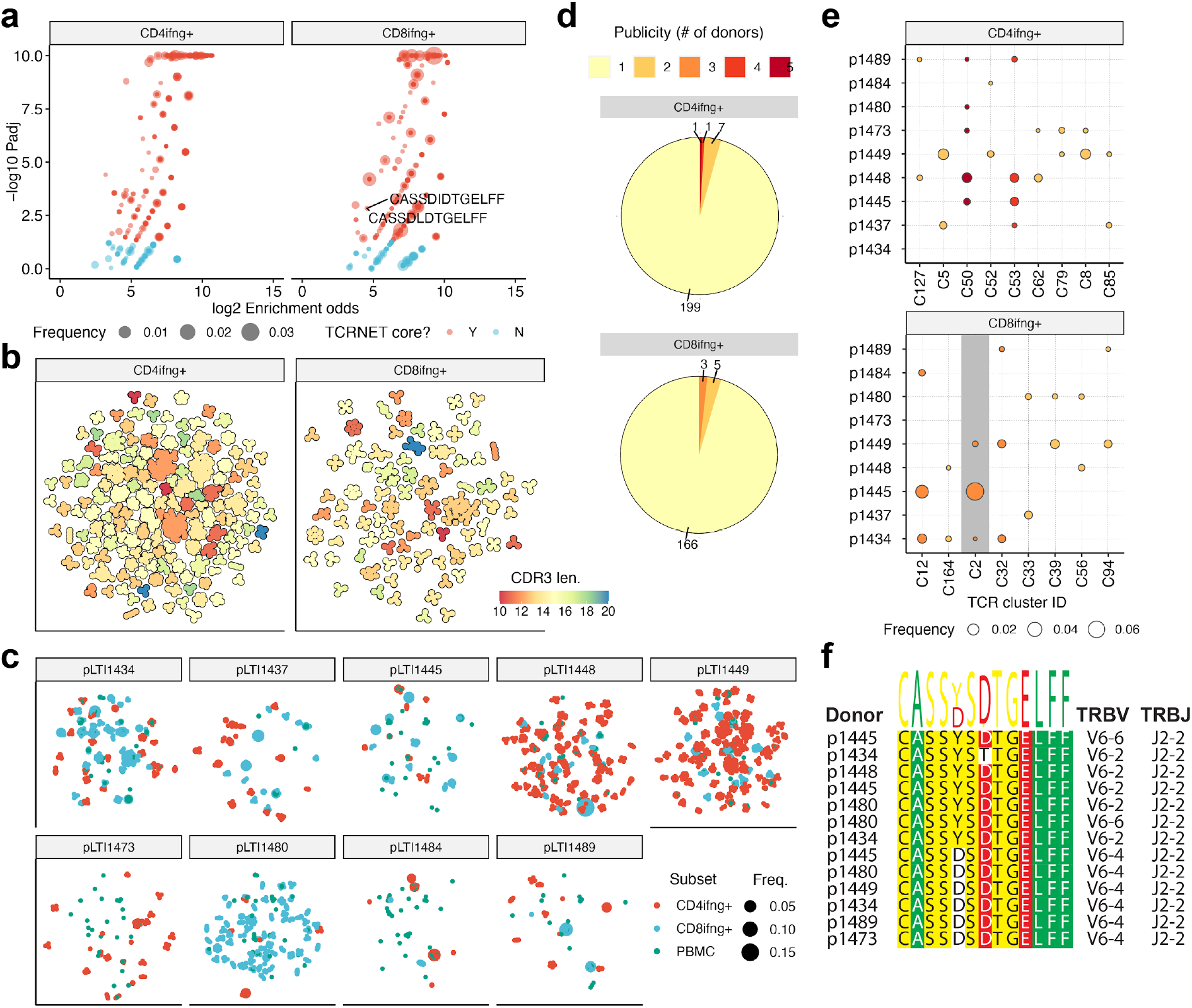
IFNγ-producing T-cells from stimulated T-cell pools feature public TCR motifs. **(a)** T-cell neighborhood enrichment (TCRNET) analysis results showing TCRs related to expanded T-cell families. The plot quantifies the number of homologous TCRs expanded upon stimulation compared to control dataset. Each point represents a TCR sequence and point size represents total frequency in a pooled sample. X axis corresponds to the logarithm of the observed to expected number of homologous TCRs (neighbours) for a given sequence, Y axis is the logarithm of adjusted P-value for TCRNET test, capped at 10^-10^. Red points specify selected TCRs (enrichment odds greater than 4x, Padj < 0.05). Labels indicate two shared TCRs found in tetramer positive T-cell dataset that are YLQ-specific. Only TCRs with Padj < 1 are shown in order to avoid overplotting. **(b)** CDR3 similarity map showing the structure of motifs discovered for CD4+ and CD8+ T-cells. Points representing individual CDR3 sequences are placed according to the layout of a TCR similarity graph, built based on CDR3 sequences identified by TCRNET. Black outline shows groups of homologous CDR3 sequences, color indicates the CDR3 region length for a given cluster. Only clusters with at least 3 members are shown. **(c)** Distribution of TCRs coming from TCRNET-identified clusters across donors. Each point represents a unique TCRs, point sizes are scaled according to TCR frequency in a given donor and subset, points are colored according to T-cell subset. **(d)** Publicity of CDR3 motifs discovered in IFNγ -producing CD8+ and CD4+ T-cell datasets. Labels indicate the number of distinct CDR3 motifs found in a given number of donors. **(e)** Distribution and frequency of TCRs associated with public CDR3 motifs in IFNγ -producing CD8+ and CD4+ T cell populations of different donors. Point size and color corresponds to a given cluster frequency in a given donor and publicity respectively (see **d**). Cluster C2 that contains TCRs found in tetramer positive T-cells (see **a**) is highlighted with grey background. Only clusters present in at least two donors are shown. **(f)** Multiple sequence alignment of the CDR3 region, Variable and Joining genes of distinct TCR variants corresponding to cluster C2 found in PBMC (unstimulated) samples from different donors.

Plotting the CDR3 similarity graph revealed many large (in terms of the number of members) homologous clusters of various CDR3 lengths, with CD4+ subset displaying higher cluster density than CD8+ (Figure 5b). The total number of clusters was higher for CD4+ subset than CD8+ (208 vs 174 clusters), the average CD4+ cluster size was larger than CD8+ in terms of number of unique CDR3 amino acid sequences (5.8 vs 4.0), yet the average frequency of a given cluster in corresponding dataset in terms of number of cells was larger for CD8+ than CD4+ (0.22% vs 0.12%), altogether highlighting higher diversity of CD4+ response in line with previous reports (Qi, Liu et al. 2014).

We then mapped the set of detected TCR clusters back to original donor samples (Figure 5c) finding that all donors feature some level of homologous TCR response, yet there are some prominent inter-donor differences: for example, donors p1448 and p1149 displayed many CD4+ clusters, while donor p1480 displayed almost exclusively CD8+ clusters, and donor p1484 displayed few clusters some of which are nevertheless of high frequency in terms of the number of cells. Analysis of cluster sharing across donors (Figure 5d) revealed that most clusters are private to donors that is expected as donors have multiple unmatched HLAs and cells are stimulated with whole protein, however, there were 9 CD4+ and 8 CD8+ clusters shared between stimulated cells of multiple donors (Figure 5e).

One of the “public” clusters (C2 in Figure 5e) contains sequences matching those found in YLQ-specific T-cells and was further explored by mapping it to unstimulated PBMC samples of all donors (Figure 5f). Exploring both stimulated IFNy-producing CD8+ cells and PBMCs revealed a set of TCR sequences in multiple donors matching those of cluster C2, all sharing the same highly conserved CDR3 motif “CASS[YD]SDTGELFF”.

## Discussion

Here we analyzed T cell and humoral immune response to SARS-COV-2 in 31 donors recently recovered from COVID-19 and two control cohorts of healthy donors sampled before and during the onset of COVID-19 pandemic. Two patients (p1472 and p1473) had no detectable antibody levels in the serum to any of the tested SARS-CoV-2 antigens and no T-cellular response to any of the peptide pools, albeit they had T-cells reactive to the recombinant S-protein. One possible explanation is the delayed seroconversion (blood was sampled at 22 and 36 days since the onset of symptoms or positive PCR test) which was described to occur as late as day 50 (Wajnberg, Mansour et al. 2020) though in another antibodies developed in 100% patients at days 17-19 (Long, Liu et al. 2020). The other possibility is the false positive PCR test. In our study as in (Phan, Subramanian et al. 2020) detection of anti-RBD IgG yielded more reliable results than other tested antigens. Previously it was demonstrated that titers of anti-SARS-CoV-2 antibodies positively correlate with patient age (Wu, Wang et al. 2020). In our cohort we did not observe that (Figure S2d-f).

The accumulating data indicate that some healthy COVID-19-naive donors have T cells specific to SARS-CoV-2 antigens and in particular S-protein (Braun, Loyal et al. 2020, Grifoni, Weiskopf et al. 2020, Ni, Ye et al. 2020). Our findings are in line with that with acaveat that we observed the significant increase of SARS-CoV-2 S-protein reactive T-cells in healthy donors sampled during the COVID-19 pandemic (Figure 2c and 2d). Combined with the complete lack of SARS-CoV-2 specific antibodies in that group this suggests that some of the donors may have had contact with SARS-CoV-2 before blood sampling. Due to cross-reactivity induced by other coronaviruses their T cells might have protected them from developing the full scale infection. This is illustrated by the case of p1477 who is known to cohabited with COVID-19 patient but was negative in multiple PCR tests, did not have any COVID-19 typical or flu-like symptoms and had no detectable antibodies to any of the SARS-CoV-2 antigens. This hypothesis, although, needs to be validated on a larger cohort of donors.

As others have shown before, we observed that significantly more CD4+ cells in convalescent donors express HLA-DR and CD38 (Braun, Loyal et al. 2020, Thevarajan, Nguyen et al. 2020). We have also shown the same tendency for CD8+ T cells, though the difference was not significant (Figure 2g). As was shown before (Weiskopf, Schmitz et al. 2020) the majority of SARS-CoV-2 specific CD4+ belonged to the T_CM_ subpopulation while CD8+ were predominantly of T_TE_ and T_EM_ phenotype.

In this work we used the IFNγ-secretion upon antigen stimulation as a criterion of antigen-specific cells. This approach might potentially miss some relevant T cells as there are other ways of T-cell reactivity, in particular for CD4+ T cells. Nevertheless, previous study showed that CD4+ T cells reacted to stimulation by SARS-CoV-2 antigens (S-protein in particular) mostly by Th1-type response (Weiskopf, Schmitz et al. 2020). In another study IFNγ was also the predominant cytokine produced by memory T cells after stimulation with SARS-CoV-2 peptides (Fan, Huang et al. 2009).

The results we obtained for T-cells stimulated by the full-length S protein reveal a highly specific response in terms of TCR repertoire structure of both CD8+ and CD4+ T cells. IFNγ-producing T-cell repertoires featured multiple groups of homologous TCR sequences that are in a good agreement with TCR variants found to be enriched based on corresponding clonal abundances. Thus, we were able to identify hundreds of TCR motifs, some of which were shared across multiple donors. Interestingly, only one of the detected motifs was found in tetramer positive CD8+ T-cell fraction, suggesting that immediate response may be targeting a wealth of distinct S-protein epitopes, some of which are presented by HLA alleles other than HLA-A*02. This single motif, however, displayed extensive sharing between donors and was also found in an unstimulated PBMC fraction.

In this work we tested 13 epitopes for 11 of which presentation by HLA-A*02 was previously confirmed and immunogenicity demonstrated in SARS-CoV convalescent patients (Zhou, Xu et al. 2006) and healthy donors (Lv, Ruan et al. 2009). Despite antigen-specific expansion being performed for all peptides to detect even the small T-cell clones, we observed consistent epitope-specific response to only 2 of 13 peptides (Figure 3a). In particular we did not see a strong response to RLN peptide despite its previously reported immunodominance in SARS-CoV-2 convalescent patients (Wang, Chen et al. 2004). KLP-specific T cells were detected only in 1 convalescent patient, contrary to study on SARS-CoV (Zhou, Xu et al. 2006), which showed this epitope to be highly immunogenic in convalescent patients but not healthy donors. KLP peptide of SARS-CoV-2 differs from SARS-CoV by a single amino acid substitution (M-T), whereas RLN peptide is 100% homologous. The lack of the immune response to the studied epitopes in SARS-CoV-2 convalescents may be attributed to the existence of more immunodominant epitopes, that elicit T cell expansion, which outcompetes RLN- and KLP-specific T cells. It was previously shown that T-cell immune response in healthy donors is focused predominantly on epitopes in C-terminal part of SARC-CoV-2 S-protein which may be explained by higher homology of this region with common cold coronaviruses (Braun, Loyal et al. 2020). In our study only one epitope (YLQ) was derived from the N-terminal part of the S-protein, and we saw almost no response to this epitope in healthy donors.

We described two HLA-A*02:01 epitopes of SARS-CoV-2 S-protein YLQ and RLQ with immune response to them detected in 13 of 14 and 10 of 14 of convalescent donors, respectively. T cells specific to both epitopes occupied only a negligible fraction of total TCR repertoire, explaining almost no intersection between tetramer-positive and IFNγ-secreting repertoires. It is possible that YLQ and RLQ-specific clones are localized in the peripheral tissues and only a limited number of cells are present in the circulation. It was previously shown that clonal CD8+ expansions in SARS-CoV-2 are tissue resident (Liao, Liu et al. 2020).

YLQ epitope was suggested before as potentially immunogenic in SARS-CoV-2 (Baruah and Bose 2020, Romero-López, M et al. 2020). It was also predicted to bind to HLA-A*01:01 and HLA-C*07:02, besides HLA-A*02:01 (Lee 2020). In this study we showed that it is not only highly immunogenic but also is recognized by T cells sharing the same TRA V-segment (TRAV12-1), suggesting the important role of the TCR alpha chain in recognition of this epitope. Of note this epitope as a substantial share of N-terminal part of S-protein was not present in the utilized peptide pool, possibly explaining the discrepancy between reactivity to the recombinant S-protein and peptide pools as seen on Figure S4.

Our analysis of T cells specific to RLQ and YLQ epitopes obtained using tetramer-based enrichment revealed a set of highly conserved TCR sequences shared across multiple donors. These sequences feature highly restricted Variable segment usage and relatively short CDR3 length, suggesting that RLQ and YLQ are targeted by public TCRs that rely on germline-encoded motifs to recognize them. The YLQ epitope is recognized by several unrelated motifs that are shared across several donors, suggesting that the response is both public and diverse.

Alongside (Minervina, Komech et al. 2020) this study provides a first glimpse into the structure of T-cell response to SARS-CoV-2. Further studies on the specificity of the SARS-CoV-2 targeted response and deconvolution of SARS-CoV-2 epitopes would provide crucial information for vaccine design and disease diagnosis.

## Data Availability

TCR sequencing data was deposited to the VDJdb database (vdjdb.cdr3.net) and attached to this article as Extended data tables

https://www.vdjdb.cdr3.net

## Acknowledgments

Authors would like to acknowledge their gratitude to all donors who volunteered for our study; to the nurses who performed the venipuncture: Yulia Fadeeva, Anastasya Nisanova, Valentyna Mirponova, and Lubov Piskunova; to our colleagues: Anastasia Minervina, Mikhail Pogorelyy, Vasily Lazarev, Maria Lagarkova, Ivan Zvyagin, Igor Fabrichny, Anastassia Semikhina, Alexey Panov, Ekaterina Avilova, Artem Demidenko, Alexander Veretennikov, Fedor Rozov, Elena Osipova, Nikolay Mugue, Igor Fabrichny, Yuri Lebedin and Ekaterina Morozova for their kind help with experiments and reagents and the most valuable advice. Mikhail Shugay was supported by RFBR grant No 19-34-70011. Alexander Ivanov was supported by the Ministry of Science and Higher Education of the Russian Federation [Agreement No. 075-15-2019-1660].

## Author Contributions

Conceptualization, G.E.; Methodology, G.E., A.S., M.V., and M.S.; Software, M.S. D.Sh., D.B., A.Piv., A.Kh. and D.M.; Formal Analysis, M.V., A.T. A.Kh. and M.S.; Investigation, A.S., M.V., K.Z. S.Sh. Y.P. Y.S., A.T., A.Kh., A.M., N.S., B.B. and E. Kh.; Resources, D.K, M.M., D.D., A.Sh., A.Pil., and A.I.,; Writing - Original Draft, G.E., A.S., and M.S.; Writing - Review & Editing, G.E., A.S.,Y.P., A.Sh, S.Sh., A.T. and M.S.; Project Administration, G.E.; Funding Acquisition, G.E., A.I. and M.S.

## Material and methods

### Donor selection

31 COVID-19 convalescent patixents from Moscow, Russia were recruited voluntarily. COVID-19 was confirmed either by positive SARS-CoV-2 RT-PCR test or retrospectively by the detection of anti-RBD antibodies. All donors signed the informed consent approved by the National Research Center for Hematology ethical committee before the enrollment. According to the classification by the US National Institutes of Health, the severity of disease was defined on patient’s case history - asymptomatic (lack of symptoms), mild severity (fever, cough, muscle pain, but without respiratory difficulty or abnormal chest imaging) and moderate/severe (lower respiratory disease at CT scan or clinical assessment, a saturation of oxygen (SaO2) >93% on room air, but lung infiltrates less than 50%). In this study were also included 7 volunteers, sampled during COVID-19 pandemic but without known contact with COVID-19 patients (except p1477, who was cohabiting with a COVID-19 patient). Additionally, 10 healthy hematopoietic stem cell donor samples and 10 healthy donor serum samples were obtained from the blood bank with the approval of the local ethical committee. Cell samples were cryopreserved no later than September 2019, serum samples no later than 2017.

### HLA genotyping

For most donors HLA genotyping was performed by next-generation sequencing using ALLType kit (One Lambda, Canoga Park, California). This kit uses a single multiplex polymerase chain reaction to amplify the full HLA-A/B/C gene sequences and from the exon 2 to the 3’UTR of the HLA-DRB1/3/4/5/DQB1 genes. Prepared libraries were run on an Illumina MiSeq sequencer using standard flow cell with 2 x 150 paired-end sequencing. Reads were analyzed using the HLA TypeStream Visual Software (TSV) (One Lambda), version 2.0.0.27232 and the IPD-IMGT/HLA database 3.39.0.0. Other donors were HLA genotyped by Sanger sequencing was performed for loci HLA-A,B,C, DRB1 and DQB1 using Protrans S4 and Protrans S3 reagents respectively. The PCR product for sequencing was prepared by BigDye Terminator v1.1. Capillary electrophoresis was performed on Genetic Analyser Nanophore05. One donor HLA genotyping was determined based on exome sequencing data.

### SARS-CoV-2 S-protein peptides

Putative HLA-A*02:01 epitopes of viral S-protein were included in the analysis if they meet the following criteria: 1) weak binders (0.5<rank<2) or strong binders (rank<0.5) according to NetMHCpan 4.0 2) full or partial homologs of existing SARS-CoV S-protein epitopes (identity >60%) (1) Detailed information about selected peptides is listed in Table 1. Predicted proteasomal cleavage score of the C-terminal amino acid was estimated using NetChop 3.1 (Nielsen, Lundegaard et al. 2005). HLA-A02:01 binding affinity score and rank were estimated by NetMHCpan 4.0 (Reynisson, Alvarez et al. 2020). SARS-CoV identity was measured as the count of identical positions in the alignment of amino acid sequences of SARS-CoV and SARS-CoV-2 (MN908947.3) S-protein performed by QIAGEN CLC Genomic Workbench software.

Peptides were synthesized by using a solid-phase synthesis method and purified by high-performance liquid chromatography (HPLC) (greater than 95% purity). All peptides were dissolved in DMSO, except cysteine-containing peptides which were dissolved in MES buffer, pH 6.5 / isopropanol mixture (1:1 vol.).

### PBMC isolation

Venous blood from healthy donors and recovered COVID-19 patients was collected into EDTA tubes and subjected to Ficoll (Paneco) density gradient centrifugation (400g, 30 min). Isolated PBMC were washed and used for multiple assays.

### Flow cytometry

A surface staining and phenotype analysis of the PBMC were performed with CD3-AF700 (OKT3; Sony), CD4-FITC (RPA-T4; Sony), CD8-PerCP/Cy5.5 (RPA-T8; Sony), CCR7 (CD197)-PE/Dazzle594 (G043H7; Sony), PD1 (CD279)-BV421 (EH12.2H7; Sony), CD27-BV711 (0323; Sony), CD28-BV785 (CD28.2; Sony) and CD45RO-PE/Cy7 (UCHL1; Sony). Cells were analyzed on the FACS

Aria III cell sorter (BD Biosciences). FlowJo Software (version 10.6.1., Tree Star, Ashland, OR) was used for analyzing data.

### Tetramer staining

Antigen-specific cells were detected by staining, with CD3-AF700 (OKT3; Sony), CD8-FITC (Sony), 7AAD (Sony) along with 7 combinations of two different peptide-tetramer complexes conjugated with Streptavidin-Allophycocyanin and Streptavidin-R-Phycoerythrin (Thermo Scientific). FACS Canto II cell analyzer and Aria III cell sorter (both BD Biosciences) were used. Data were analyzed using FlowJo Software.

### IFNγ secretion assay

Measurement of IFNγ secretion in CD4+ and CD8+ T cells was performed using IFN-γ Secretion Assay-Detection Kit (APC) (Miltenyi Biotec) according to the manufacturer’s protocol. Briefly, fresh PBMC isolated from biobanked healthy donors (BB), sampled during COVID-19 pandemic and COVID-19 convalescent patients (CP) were resuspended in RPMI 1640 culture medium (Gibco) supplemented with 5% normal human A/B serum (obtained from pooled inactivated human AB Rh-male serum) and 1mM sodium pyruvate (Gibco) and plated at density 1-10×10^6^ cells/ml. Cells were treated for 16h with 10μg/mL of SARS-CoV-2 S-protein, followed by incubation for 5 min at 4°C with IFNγ Catchmatrix Reagent (Miltenyi Biotec). Cells were then transferred into a warm medium (37°C) for 45 min to re-initiate secretion of IFNγ, washed and stained with surface and phenotype markers together with IFNγ Detection Antibody-APC (Miltenyi Biotec) for 10 min at 4°C. CD4+IFNγ+ and CD8+IFNγ+ populations were sorted directly to TRIzol™ Reagent (Thermo Fisher Scientific) using FACS Aria III cell sorter (BD Biosciences). Data were analyzed using FlowJo Software (version 10.6.1., Tree Star, Ashland, OR). For group comparison we used Kruskal-Wallis test and Dunn’s multiple comparison test and Mann-Whitney test.

### Immunomagnetic isolation of CD4+ and CD8+ T-cells

CD4+ and CD8+ T-cells were isolated using human CD4+ MicroBeads (Miltenyi Biotec) and human CD8+ MicroBeads (Miltenyi Biotec), respectively, according to the manufacturer’s protocol. Briefly, the PBMC isolated from COVID-19 convalescent donors (CD), were incubated for 15 min at 4°C with lyophilized CD8+ MicroBeads, washed and loaded onto MS MACS Column (Miltenyi Biotec), which were placed in MACS Separator. After columns were removed and magnetically labeled CD8+ cells were eluted, the flow-through fraction was collected and used for isolation of CD4+ cells by lyophilized CD4+ MicroBeads. Isolated CD8+, CD4+ T-cells, and unlabeled cells (source of APC) were counted and used for IFNγ ELISPOT assay.

### IFNγ ELISPOT assay

Measurement of antigen-specific IFNγ production by T cells was performed using ImmunoSpot Human IFN-γ Single-Color ELISPOT kit (CtL, HIFNγ P-2M/5) with 96-well precoated with human IFNγ capture antibody nitrocellulose plate. CD8+ and CD4+ T cells isolated with the use of immunomagnetic beads were plated at density 10^5^ cells/well in duplicate. Unlabeled cells obtained after the selection of CD8+ and CD4+ T cells were used as APC at density 2×10^5^/well. SARS-CoV-2 S-protein was added at a final concentration of 10μg/mL in Serum-free Testing Medium (CTL) containing 1mM GlutaMAX (Gibco) at a final volume 200μL/well. Total PBMC were seeded at a concentration 5×105/well in duplicates and pulsed with M-, N- or S peptide pools (Miltenyi Biotec) at a final concentration 1μM. Plates were incubated for 18h at 37°C in 9% CO2. After, plates were washed two times with PBS and then two times with PBS, containing 0.05% Tween-20, followed by incubation with biotinylated anti-human IFN-γ Detection antibody for 2h at room temperature (RT). Wells were washed three times with 0.05% Tween-20/PBS and Streptavidin-AP were added for 30 min at RT. After a few washings, the colorimetric reaction was started by adding substrate components for 15 min at RT. The reaction was stopped by gently rinsing the plate with tap water. Spots numbers were counted by CTL ImmunoSpot® Analyzer using ImmunoSpot® Software. For group comparison the data were log(2)-transformed, the normality was assessed by Shapiro-Wilk test and two-way ANOVA with Tukey’s multiple comparisons test was performed.

### Expression and purification of recombinant proteins

The recombinant S-protein-His6 of SARS-CoV-2 was encoded by the plasmid kindly provided by prof. Florian Krammer (7) was expressed in the Expi293 Expression System (ThermoFisher Scientific) for five days. After harvesting the medium was centrifuged at 10000g, the supernatant was concentrated 10 times and diafiltered into buffer A (10 mM phosphate buffer, 2,7 mM KCl, 500 mM NaCl, pH 8.0) using ÄKTA™ flux tangential flow filtration system (Cytiva, filter cartridge UFP-10-C-4X2MA (cat. # 56-4102-11)). The concentrate was mixed with Ni-NTA agarose resin (Qiagen) previously equilibrated with buffer A and incubated 2h at 22°C with agitation. The resin mix was packed into a column and washed with 10 volumes of buffer A with 30 mM imidazole and eluted with buffer A with 200 mM imidazole. The eluate was dialyzed against 100 volumes of PBS (10 mM phosphate buffer, 2,7 mM KCl, 137 mM NaCl, pH 7,5) using Slide-A-Lyzer^TM^ Dialysis cassettes (20K MWCO, Thermo Fisher Scientific).

Biotinylated MHC class I/UV-cleavable peptide complexes for UV-mediated ligand exchange were produced as described (Rodenko, Toebes et al. 2006, Bakker, Hoppes et al. 2008). Briefly, heavy (HLA-A*02:01 with biotinylation tag) and light (beta-2 microglobulin) chains were expressed in *Escherichia coli* strain BL21(DE3) pLysS in the form of inclusion bodies. Proteins were dissolved in the denaturation buffer (50 mM Tris-HCl, pH 8.0, and 8 M urea). In vitro folding was set up in folding buffer (100 mM Tris-HCl, 400 mM arginine, 5 mM reduced glutathione, 0.5 mM oxidized glutathione, 2 mM EDTA, protease inhibitors, 1 mM PMSF, pH = 8.0). UV-cleavable peptide (KILGFVFJV for HLA-A*02:01 and AARGJTLAM for HLA-B*07:02, Thermo Scientific custom peptide synthesis), light and heavy chains were mixed in the folding buffer at the 30: 2: 3 molar ratio. Correctly folded complexes were purified on Superdex 75 pg column (Cytiva) using tris-buffered saline (20 mM Tris-HCl, 150 mM NaCl, pH 8.0) as mobile phase. Complexes were biotinylated by in-house made biotin ligase and purified on Superdex 75 pg columns.

Concentrations were determined using specific molar absorption coefficient /A1cm0.1%280 = 1.03, 2.36, and 1.68 for S-protein-His6, HLA-A, and hB2M, respectively (calculated in SnapGene® Viewer based on amino acid sequence).

The recombinant SARS-CoV-2 N-protein was a generous gift from Vasily Lazarev.

### ELISA

IVD ELISA kit developed by the National Research Center for hematology was used for the detection of anti-RBD IgG according to the manufacturers’ instructions. For detection antibodies to N-protein and full-length S-protein, we used in-house ELISA assay according to the protocol adapted from Kramer et al. (7). In brief, 96 well plates (Thermo Scientific, cat. 9502227) were coated with 50 μL per well of 1.4 μg/mL solution of N-protein or 0.4 μg/mL solution of S-protein. The proteins were diluted in a coating buffer (bicarbonate/carbonate 100 mM, pH 9.6). 14h later the plates were washed 3 times with 250 μL of PBS with 0.1% Tween 20 (TPBS) and blocked with 200 ul of 3% non-fat dry milk (Thermo) in PBS for 1.5h. Then the plates were washed thrice and 100 ul of serum samples diluted (1:100) in 1% non-fat dry milk prepared in TPBS were added in duplicates and incubated for 2h. Next, the wells were washed 3 times and were incubated for one more hour with 100 μL of anti-human IgG monoclonal HRP-conjugated antibodies (supplied with RBD ELISA Kit). Finally, the plates were washed 3 times and 100 μL of 3′,5,5′-Tetramethylbenzidine (TMB) substrate were added to each well. 10 min after 50 ul of 1 M H3PO4 was added as a stop-solution and optical density (OD) was measured at 450 nm with a reference of 650 nm.

To compare the samples within one plate, as well as normalize the values across the different plates and compare them, we performed the serial dilutions of p1484 serum (from 1:200 to 1:51200) in each plate. A sigmoid four-parameter logistic (4PL) fitting curves model was used to fit the calibration curve based on the serial dilution. For each plate, an EC50 value (the half OD between the top and bottom segment of a curve) was used as a coefficient of normalization. The mean of two OD values for each sample was divided by the coefficient of normalization for the given plate. For group comparison Kruskal-Wallis test and Dunn’s multiple comparison test was used.

### Antigen-specific T-cell expansions

PBMC of HLA-A*02:01 positive donors were used for rapid in vitro expansion. Briefly, 3×10^6^ cells were incubated for 8-12 days in RPMI 1640 culture medium supplemented with 10% normal human A/B serum, 1mM sodium pyruvate, IL-7 (25ng/mL), IL-15 (40ng/mL), and IL-2 (50ng/mL) at final volume 2ml/well. Half of the medium was replaced on day 3, 5, and 7. A mix of HLA-A*02:01-restricted peptides (see Table 1) of S-protein in dMsO or MES buffer (Sigma-Aldrich) (final concentration of each in medium 10ng/mL) was added at day 0.

### TCR repertoire sequencing

TCR libraries were processed as described previously (Zvyagin, Mamedov et al. 2017). cDNA synthesis reaction for alpha and beta chains of T cell receptor was carried out with primer to C-terminal region and SMART-Mk, providing 5’ template switch effect and containing sample barcode for contamination control and unique molecular identifier

TCR repertoire sequencing data was analyzed using MIXCR software with default settings. Tetramer-positive TCR sequencing data was formatted and deposited to the VDJdb database (vdjdb.cdr3.net).

### TCR repertoire motif discovery and motif analysis

Motif discovery for TCR repertoires corresponding to tetramer-positive TCRs specific to SARS-CoV-2 epitopes was performed as described previously (Bagaev, Vroomans et al. 2020). Briefly, the TCR similarity network was constructed allowing a single amino acid substitution in CDR3 sequence (Hamming distance of 1), the number of similar sequences (“neighbors”) for each CDR3 was counted and compared to the expected number of neighbors predicted using a reference dataset containing ~10^7^ amino acid sequences of either TCR alpha and beta chains. CDR3 sequences having more neighbors than would be expected at random were considered to be representative (“core”) sequences, TCR motifs were defined as connected components of TCR similarity network containing at least one core sequence.

A similar analysis was performed to detect TCR motifs in pooled IFNγ+ fractions of stimulated CD4+ and CD8+ T cells, except for using a control made from pooled PBMC repertoire of corresponding donors and counting neighbors based on CDR3 nucleotide sequences as described in (Pogorelyy, Fedorova et al. 2018).

R markdown notebooks used for data analysis are available at https://github.com/antigenomics/covid19-tcr-analysis.

## Supplementary information

**Figure S1.**
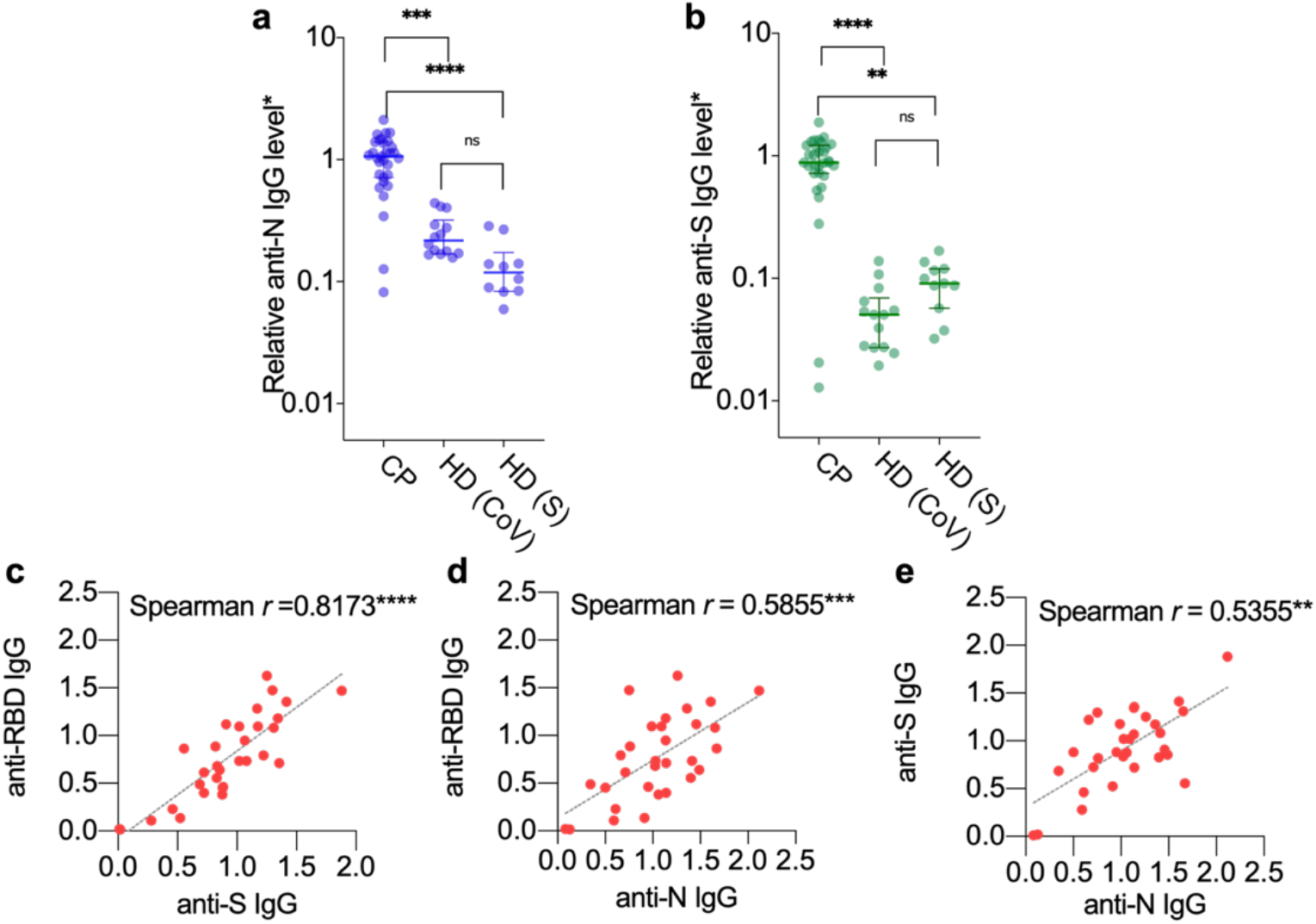
Antibody response to the different SARS-CoV-2 antigens. For detection antibodies to N-protein **(a)** and full-length S-protein **(b)** in-house ELISA assay was used. OD650 was subtracted from OD450 for each well. Mean OD for each serum sample was divided to normalising coefficient (EC50 of the calibration curve) in order to compare the samples across different plates. The Spearman correlation and linear regression between anti-RBD and anti-S IgG **(c)**, anti-RBD and anti-N IgG **(d)**, as well as anti-S and anti-N IgG **(e)** is shown. For group comparison Kruskal-Wallis test and Dunn’s multiple comparison test was used. RBD - receptor binding domains CP - convalescent patients HD (CoV) - healthy donors sampled during COVID-19 pandemic, and HD (S) - biobanked healthy donors of serum

**Figure S2.**
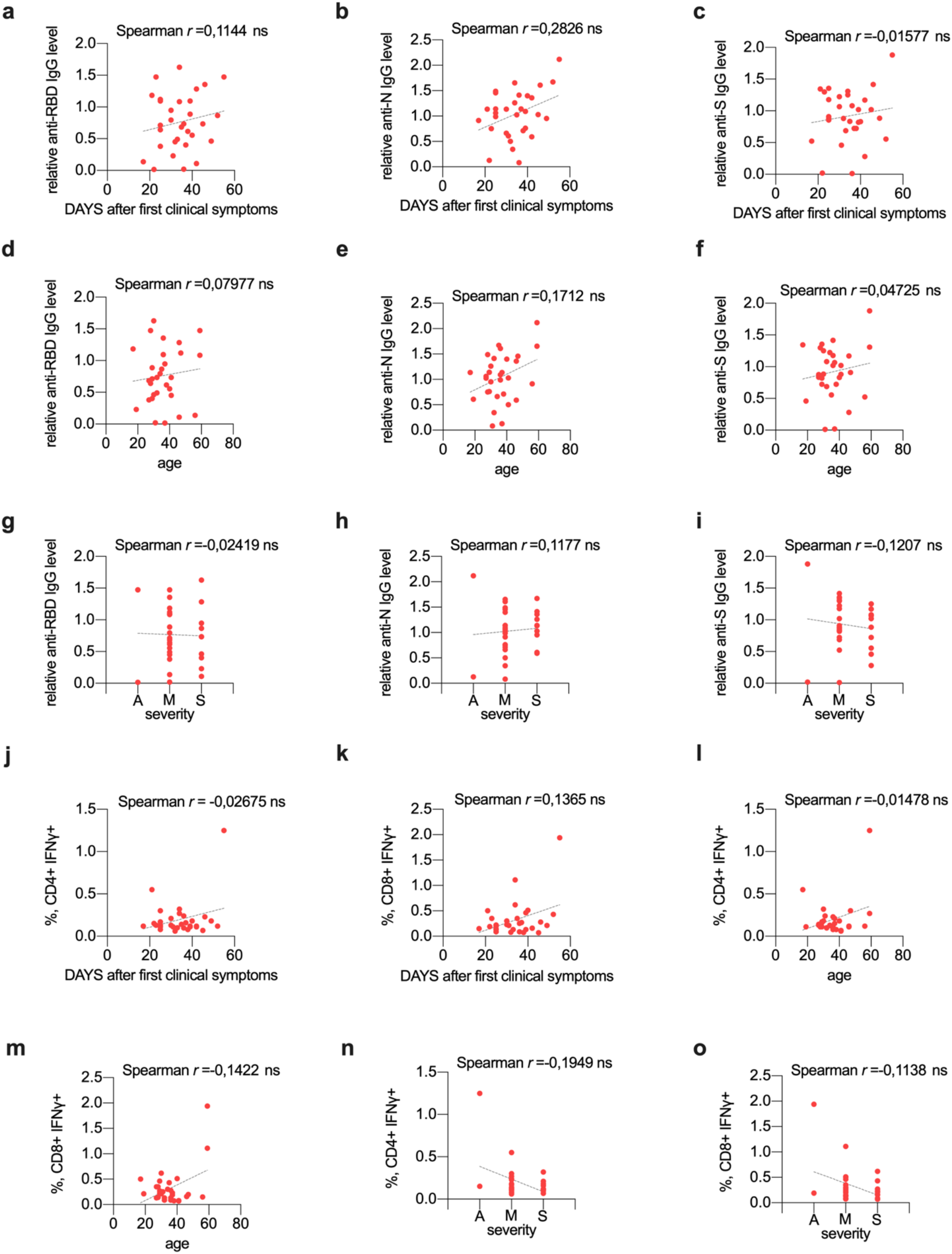
Correlation between clinical data and cellular and antibody response in convalescent donors. **(a-c)** Correlation between antibody response and time after the onset of the disease or positive PCR-test. Spearman’s coefficient of correlation between time and relative levels of anti-RBD **(a)**, anti-N **(b)**, anti-S **(c)** IgGs and linear regression was plotted (n=31). **(d-f)** Correlation between antibody response and age of convalescent patient. Spearman’s coefficient of correlation between age and relative levels of anti-RBD **(d)**, anti-N **(e)**, anti-S **(f)** IgGs and linear regression was plotted (n=31). **(g-i)** Correlation between antibody response and severity of disease of convalescent patient. Spearman’s coefficient of correlation between asymptomatic (A), mild (M) and moderate/severe (S) groups of CP and relative levels of anti-RBD **(g)** anti-N **(h)**, anti-S **(i)** IgGs, linear regression was plotted (n=31). **(j-k)** Correlation between T cell response and time after the onset of the disease or positive PCR-test. Spearman’s coefficient of correlation between time and frequencies of IFNγ producing CD4 **(j)** or CD8 **(k)** and time, linear regression was plotted (n=31). **(l-m)** Correlation between T cell response and age. Spearman’s coefficient of correlation between age and frequencies of IFNγ producing CD4 **(l)** or CD8 **(m)** and time, linear regression was plotted (n=31). **(n-o)** Correlation between antibody response and severity of disease of convalescent patients. Spearman’s coefficient of correlation between asymptomatic (A), mild (M) and moderate/severe (S) groups of CP and frequencies of IFNγ producing CD4 **(n)** or CD8 **(o)** and time, linear regression was plotted (n=31). Clinical significance p-value *p<0.05, **p<0.01, ***p<0.001, ****p<0.0001.

**Figure S3.**
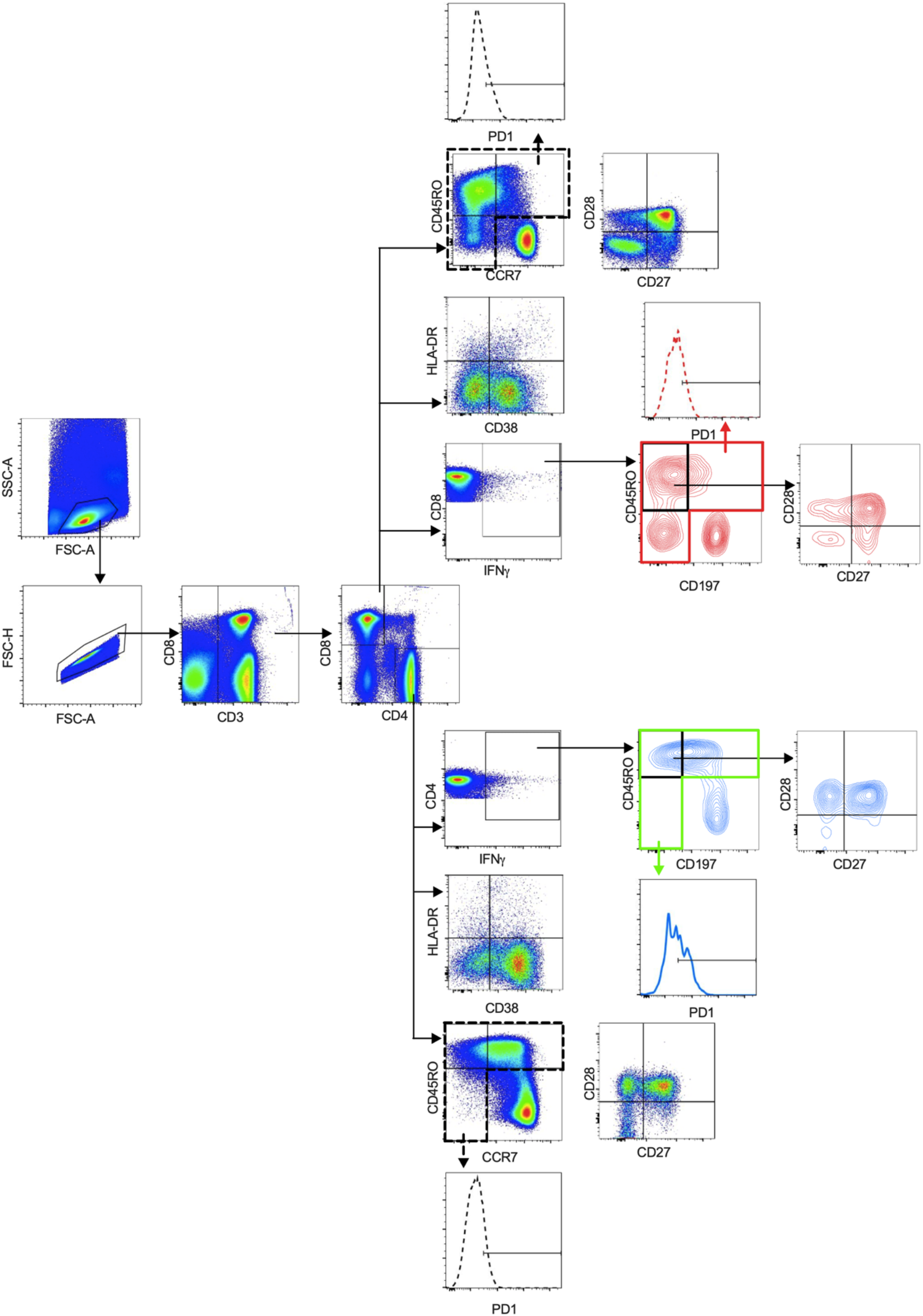
Flow cytometry gating strategy.

**Figure S4.**
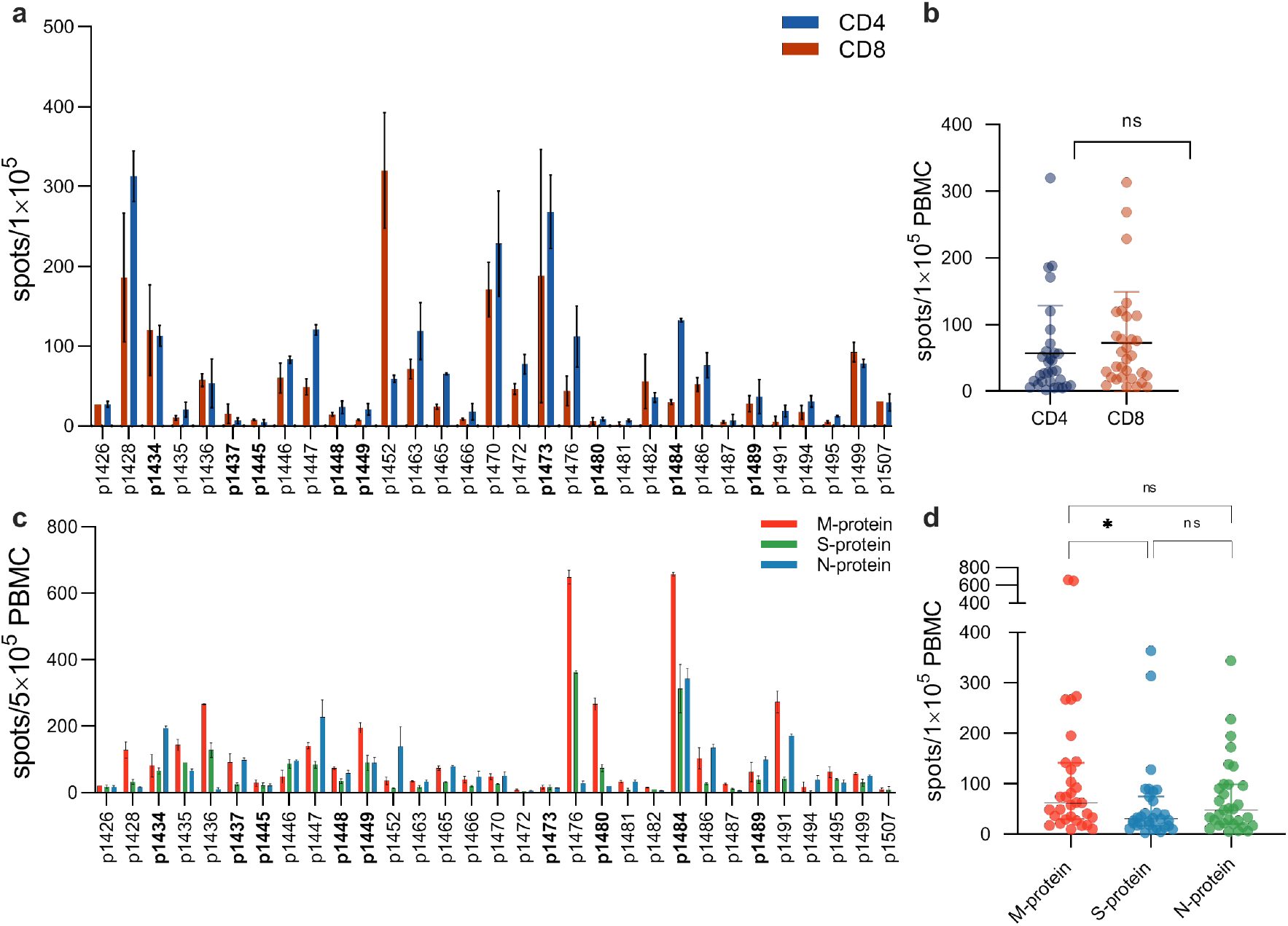
Variability of T-cell response to SARS-CoV-2 antigens. Magnetically separated CD8+ and CD4+ cells and total PBMC, isolated from COVID-19 convalescent donors (n=31) were stimulated with recombinant glycoprotein S or with pools of peptides (M, N and S) for 18 hours, respectively. INFγ response was assessed by ELISPOT. **(a-b)** Number of antigen-specific CD8+ and CD4+ T-cells. **(c-d)** Number of antigen-specific T-cells. Spots were quantified by automated digital image analysis in duplicate wells. Donors used for TCR repertoire analysis are shown in bold. For group comparison the data were log(2)-transformed, the normality was assessed by Shapiro-Wilk test and two-way ANOVA with Tukey’s multiple comparisons test was performed.*p<0.05

**Figure S5.**
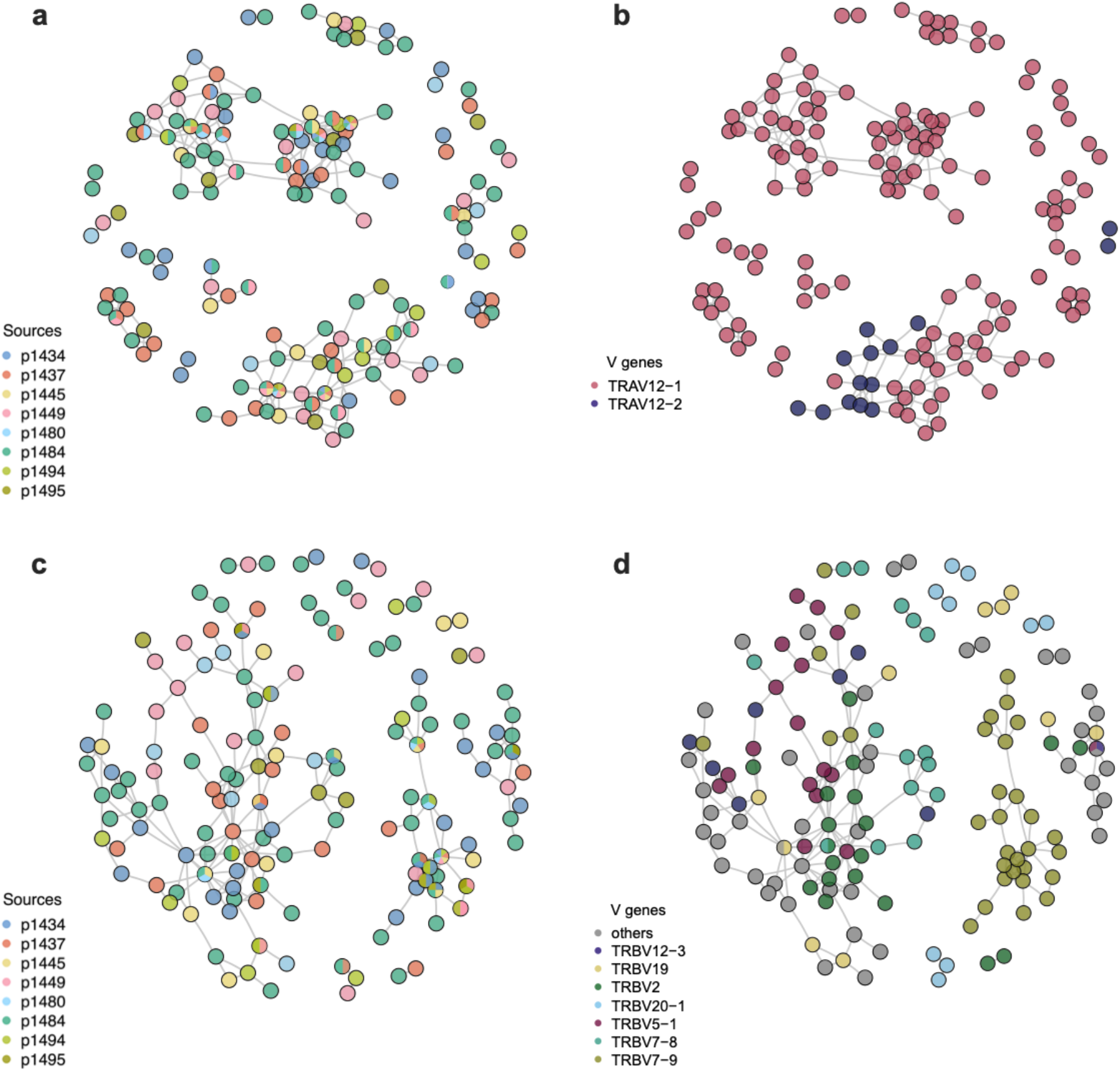
Global similarity of YLQ-specific CDR3 amino acid sequences. Graph shows CDR3 amino acid sequences of MHC-tetramer-positive clones with Hamming distance between sequences 1 or 0. **(a-b)** TCRa CDR3 amino acid sequences. **(c-d)** TCRβ CDR3 amino acid sequences. **(a-c)** Colors correspond to different CP. **(b-d)** Color corresponds to different V-genes. Each dot represents one CDR3 amino acid sequence.

**Table S1.**
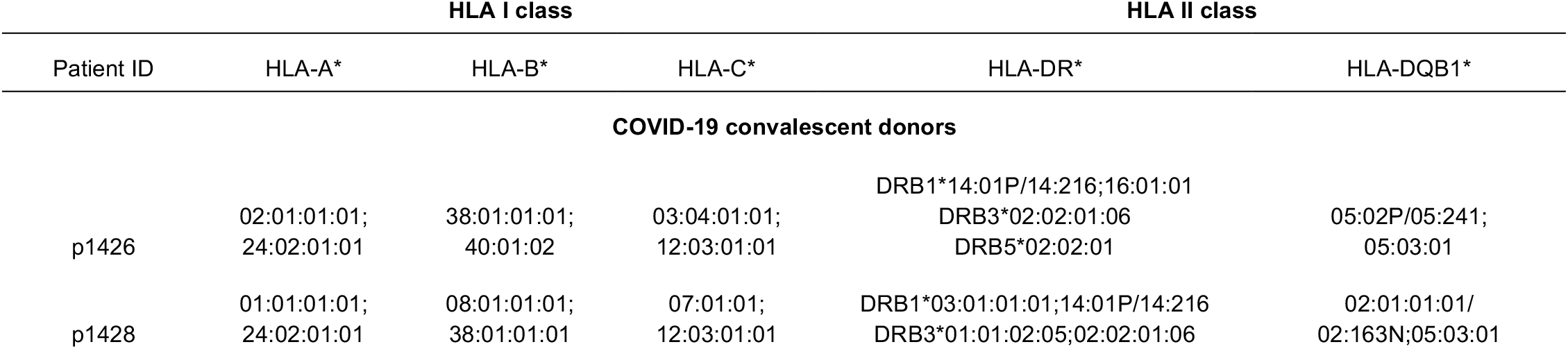
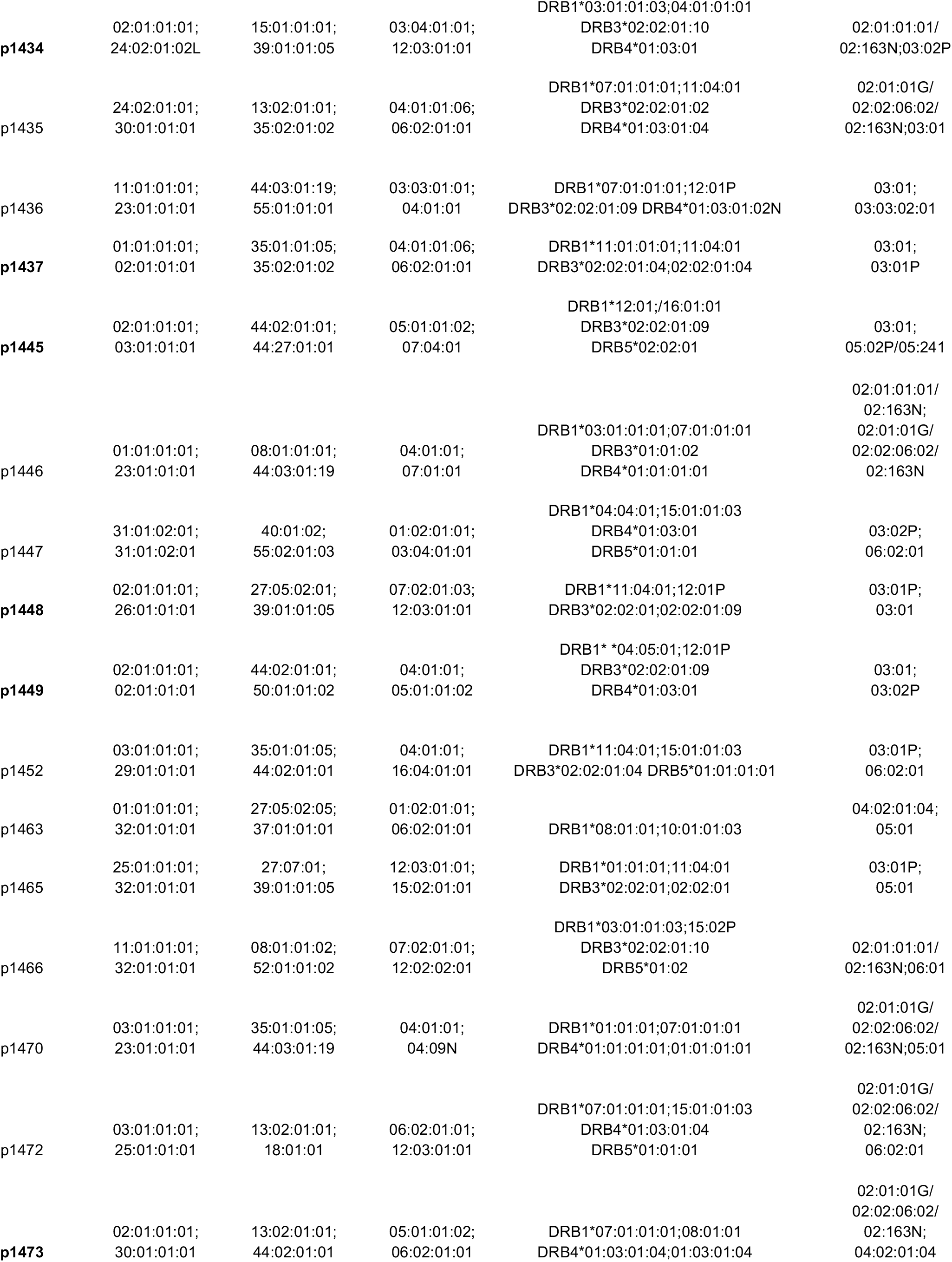
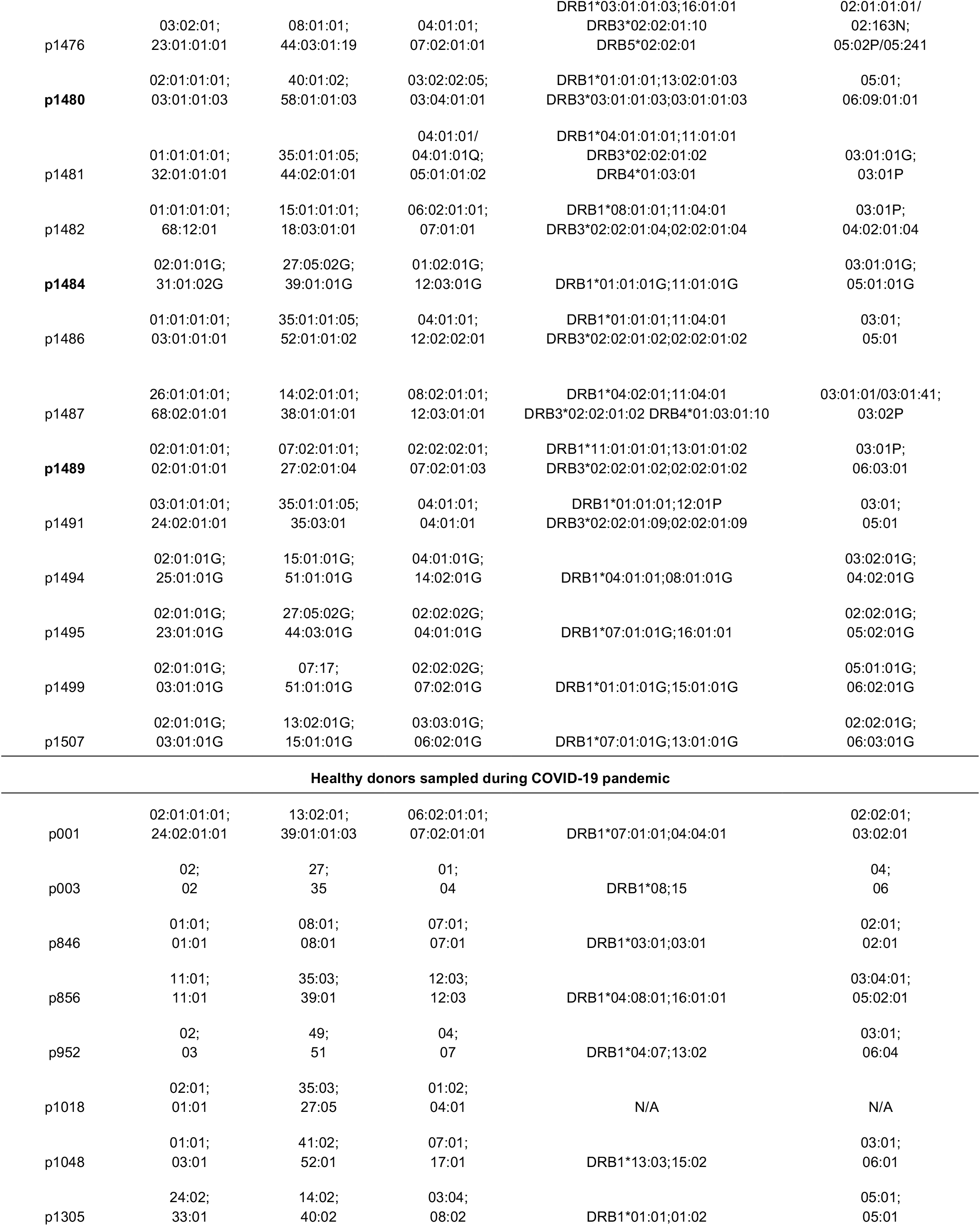
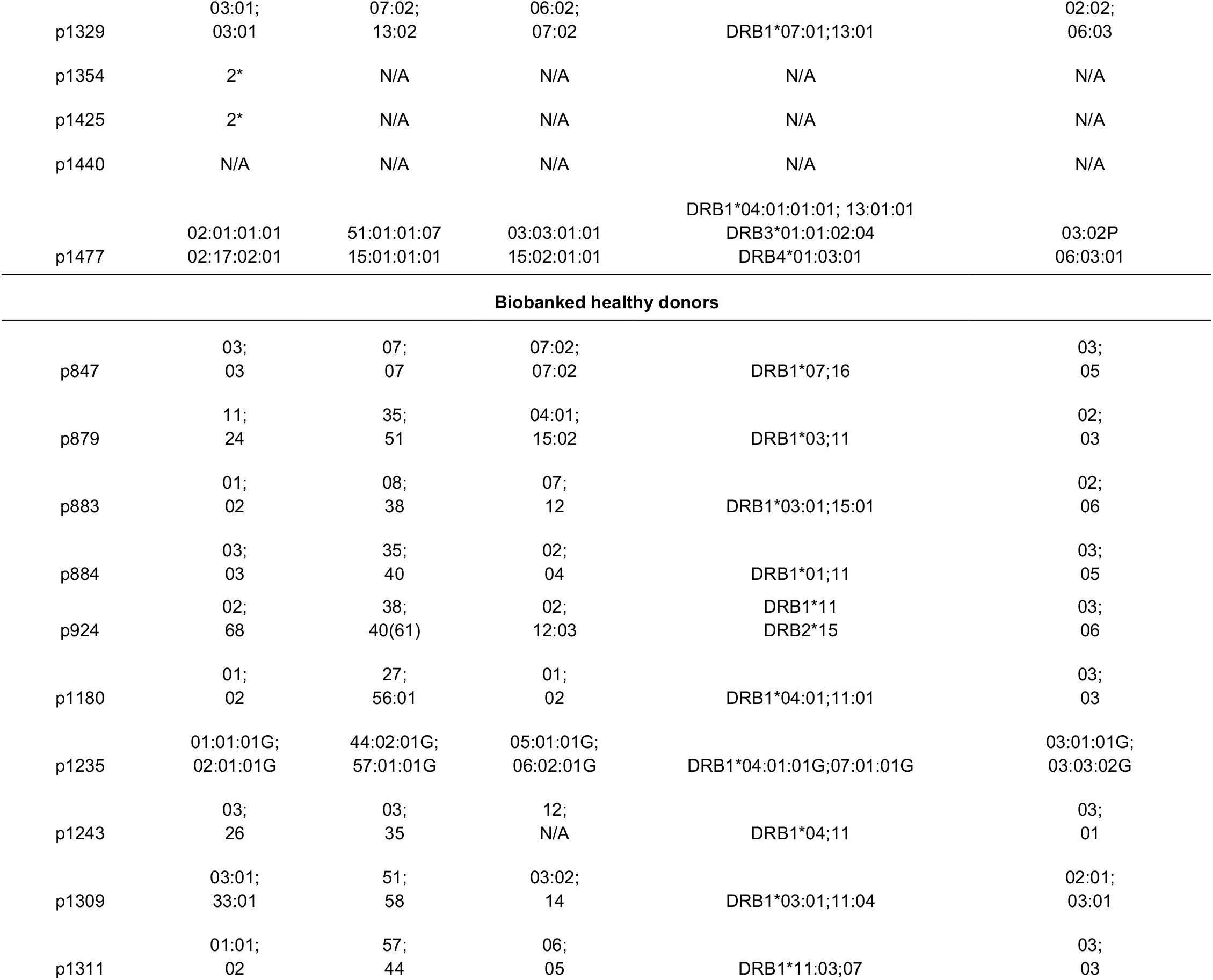
HLA genotyping (Donors for which TCR sequencing was performed are shown in bold; donors HLA-typed by flow cytometry)

**Table S2. Enriched IFNγ+ clones**

**Table S3. Homologous TCR clusters**

